# A nanopathology pipeline for clinical research across scales using human tissue

**DOI:** 10.1101/2025.08.29.25334660

**Authors:** Alana Burrell, Matthew Lawson, Paolo Ronchi, Xiaofan Xiong, Jonas Albers, Adam McLean, Michelle Willicombe, Linda Moran, Jennifer Hounsome, Martin Jones, Graham Ross, Joost de Folter, Matthew Hartley, Virginie Uhlmann, Amy Strange, Yannick Schwab, Elizabeth Duke, Candice Roufosse, Lucy Collinson

**Author notes:** These authors contributed equally to the work (Alana Burrell, Matthew Lawson, Paolo Ronchi, Xiaofan Xiong).

## Abstract

Most human tissue collected in clinic for diagnosis of pathology is formalin fixed and paraffin embedded or snap frozen, both of which destroy ultrastructure, making them unsuitable for clinical research requiring high resolution imaging. For diagnosis of some pathologies, most commonly renal and ciliopathies, a portion of the tissue is preserved optimally for ultrastructural imaging using electron microscopy (EM), but molecular antigenicity is masked. To resolve this incompatibility, we propose a protocol for fixation of human tissue in the clinic in EM-grade formaldehyde in phosphate buffer, which stabilises tissue for at least a year and preserves both ultrastructure and molecular antigenicity. To leverage this tissue for clinical research, we developed a new ‘nanopathology’ pipeline and applied it to study intravascular immune cells in kidney biopsies from patients with a transplant. In contrast to routine diagnostic practice, which images only a few percent of the whole biopsy in 2D, the nanopathology pipeline images the microstructure of the entire biopsy with X-rays using high throughput tomography (HiTT), assigns identity to immune cells using non-permeabilisation immunolabelling and confocal fluorescence microscopy of 60 µm-thick slices in regions of interest (ROIs; glomeruli and peritubular capillaries), and then images those ROIs using volume EM, specifically serial block face scanning EM. Multimodal registration of all imaging datasets into the same virtual 3D space enables extraction of nanoscale features from patient tissue that had previously only been observed in in vitro or animal models. The established nanopathology pipeline could be optimised for application to clinical research investigating a range of pathologies in the future, and with sufficient speed-up could also find application in next generation diagnostic pathways.

## Introduction

Human diseases are often diagnosed with the help of radiological investigations, molecular testing and/ or microscopy of tissue samples (histopathology/cellular pathology).

In histopathology, light microscopes are used to image tissue structure, cell morphology and nuclear morphology in stained formalin-fixed, paraffin embedded (FFPE) tissue slices. This is often supplemented with protein and/or RNA labelling to identify cell types and/or to detect pathological proteins and gene expression profiles. Histopathological diagnosis may also more rarely require examination of frozen tissue samples by immunofluorescence.

Sample preparation for FFPE and freezing tissue without strong cryoprotectants results in damage to cell membranes and organelles at the subcellular (ultrastructural) level. For the subset of pathologies where diagnostic features are too small to be seen by light microscopy, part of the tissue must therefore be processed using techniques that preserve membranes for imaging in the transmission electron microscope (TEM). Renal diseases and ciliopathies currently make up the majority of pathologies that require EM for diagnosis.

Diagnostic light and electron microscopy tend to involve imaging of 2D tissue sections, which misses 3D context and may make diagnosis challenging if the pathological change is small and/ or rare within the volume of tissue collected for diagnosis.

Meanwhile, in bioscience research, a suite of novel 2D and 3D tissue imaging and molecular phenotyping techniques (super-resolution light microscopy, spatial proteomics, spatial transcriptomics, X-ray microscopy of soft tissues, volume EM, etc) are adding new levels of structural and molecular data across scales. When combined into correlative and multimodal imaging (CMI) workflows, biological events (common and rare) can be tracked, imaged and quantified across scales, revealing the molecular and structural changes involved in pathological states in the context of whole cells, organs and organisms. Examples of new insights into biological systems gained from these novel technologies and correlative workflows include: Changes in the ultrastructure of the plasma membrane in atherosclerotic plaques in ApoE knockout mice (Hekking et al., 2009); the connectivity of projection neurons in the songbird brain (Oberti et al., 2010); the interaction of tumour cells with the endothelium during metastasis in mice (Karreman et al., 2016); ultrastructural characterisation of Lewy pathology in post-mortem human brain in Parkinson’s disease (Shahmoradian et al., 2019); and the migration of neutrophils across the blood vessel wall during inflammation in mouse muscle (Reglero-Real et al., 2021).

However, application of these novel imaging techniques and correlative workflows to clinical research using human tissue has been limited. Reasons for this include the lack of suitably preserved human tissue and the lack of genetic probes (such as GFP) to label pathological structures or events in humans. To be used in clinical research to reveal new insights into disease initiation and progression, new strategies must therefore be developed for application of these workflows to human tissue, and the following challenges must be addressed:

1) Introduce fixation procedures compatible with ultrastructural and molecular preservation into the clinic, as part of a modified diagnostic pathway that does not negatively impact on patient diagnosis.
2) Identify an imaging modality that can image cm and mm scale tissue samples with micron resolution to map tissue architecture and identify pathological regions for downstream molecular and ultrastructural imaging.
3) Develop a molecular imaging strategy that preserves membrane integrity whilst allowing penetration of probes to locate pathological structures and events, using 3D light microscopy to image the sample with the resolution to detect individual cells.
4) Develop a strategy that allows targeted 3D ultrastructural imaging of pathological regions to reveal changes at the cell, organelle and membrane level.
5) Register the multiscale, multi-resolution image datasets to interrogate ultrastructural and molecular changes in the context of the whole human tissue sample (nanopathology).

Based on previous experience in developing novel multiscale 3D imaging modalities and CMI workflows for research involving tissue from model organisms (Musser et al., 2021; Peddie et al., 2022; Vergara et al., 2021), we propose a new nanopathology pipeline for clinical research, with adaptations at each step for working with human tissue. The pipeline includes: A protocol for fixation of human biopsies in the clinic, compatible with molecular and ultrastructural preservation; a fast, high-resolution X-ray imaging workflow for fixed hydrated tissue; a targeting, sampling, labelling and molecular imaging strategy compatible with membrane preservation; targeted ultrastructural imaging of pathological regions identified by X-ray and light microscopy using volume electron microscopy; and integration of multiscale image datasets to enable interactive analysis and visualisation of pathological regions to extract novel insights into disease progression.

We illustrate the pathophysiological insights this pipeline delivers when applied to a set of human kidney transplant biopsies to investigate immune cells in the microvasculature.

## Results

### Overview of the nanopathology pipeline

The nanopathology pipeline was developed to investigate transplant rejection, with a focus on antibody-mediated rejection (AMR). In AMR, antibodies against mismatched proteins between the organ donor and recipient (usually against mismatched HLA antigens) deposit on the endothelium of the kidney microvasculature, which comprises the small capillaries of the glomeruli and the interconnecting capillary network between the tubules (peritubular capillaries - PTCs). The aim was therefore to detect: 1) the capillary network (glomeruli and PTCs); and 2) immune cells within the capillaries. For pipeline development and refinement, we used three human kidney transplant biopsies without AMR first. The pipeline is illustrated in Figure 1 and further detailed in paragraphs below.

**Figure 1.**
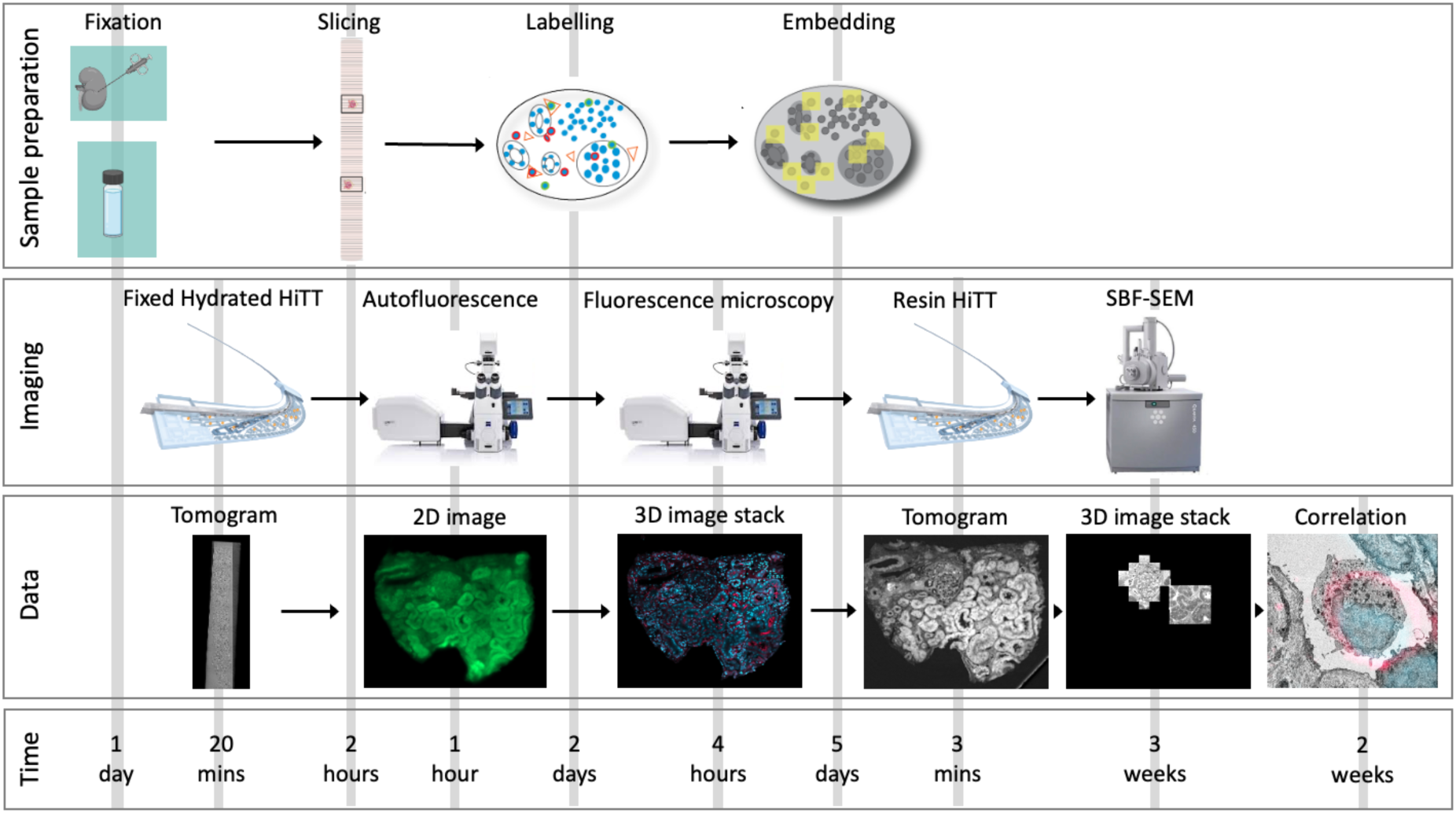
Overview of the nanopathology pipeline. A portion of the patient kidney biopsies taken for diagnosis of transplant rejection were fixed in 4% formaldehyde in 0.1 M PB to maintain compatibility with downstream molecular and ultrastructural analysis. Imaging the entire biopsy using volume EM would take around 50 years, so to reduce this time the biopsies were first shipped to the P14 synchrotron beamline for High Throughput Tomography (HiTT). HiTT revealed the microscale structure of the tissues and enabled downstream selection of regions of interest (ROIs; glomeruli and peritubular capilleries) for targeted molecular and ultrastructural imaging. Target ROIs were then cut into 60 µm thick slices using a vibrating blade microtome (vibratome), and slices imaged using autofluorescence to confirm the tissue content, and using confocal fluorescence after labelling with CD16 (for immune cells) and Hoechst (for cell nuclei). Slices were embedded for volume electron microscopy, and reimaged using HiTT, which supported high resolution targeting for imaging using serial block face scanning electron microscopy. All image datasets were registered in the MoBIE software and ROIs containing immune cells were extracted for qualitative analysis. Approximate timings for each step in the pipeline are shown.

### Collecting biopsy material compatible with the nanopathology pipeline

In routine clinical practice, a portion of the kidney transplant biopsy is fixed in glutaraldehyde, stained with heavy metals, dehydrated, and infiltrated with epoxy resin that is then polymerised to form a resin block. This procedure preserves and stabilises the tissue ultrastructure so that the sample can be used for diagnostic EM if clinically indicated. However, the procedure destroys or masks antigens, precluding downstream labelling for molecular content.

To preserve the ultrastructure and molecular antigenicity of the biopsy tissue in a format compatible with clinical research as well as diagnostic EM, a modification to the diagnostic pipeline was introduced (Supp.Fig.1). A piece of the biopsy, generally between 3 and 10 mm long, that would have been fixed using glutaraldehyde for diagnostic EM was instead fixed in 4% EM-grade formaldehyde (FA) in 0.1 M phosphate buffer (PB), before transferring it to the local diagnostic EM facility and storing in the same fixative solution at 4°C. If needed for diagnosis, the sample could be transferred to glutaraldehyde and processed as usual. If not needed for diagnosis, the biopsy tissue could be released into the nanopathology pipeline, at which point the 4% FA in 0.1 M PB was replaced with 1% FA in 0.1 M PB for long term storage at 4°C. Of the biopsy tissue collected and stored for up to 1 year before analysis, no significant degradation of ultrastructure or molecular antigenicity was observed.

### Sample selection and clinical metadata

Detailed clinical and histopathological data associated with the three kidney biopsies that were processed through the entire nanopathology pipeline were recorded (Table 1). All three patients were recipients of a deceased donor kidney transplant and received induction therapy (basiliximab or alemtuzumab) followed by baseline tacrolimus, with or without mycophenolate mofetil (MMF). All three biopsies were taken as part of a diagnostic procedure to establish the cause of graft dysfunction (i.e. a rise in creatinine). Whereas biopsies 1 and 3 were taken within about 4 months post-transplant, biopsy 2 was taken at around 6 years post-transplant. All three patients were devoid of circulating anti-donor antibodies.

**Table 1.** Clinical and diagnostic histopathological data.

|  | Patient 1<br>(EM4652) | Patient 2<br>(EM4654) | Patient 3<br>(EM4629) |
| --- | --- | --- | --- |
| Demographics and clinical features |  |  |  |
| Type of transplant | DBD | DBD | DCD |
| CIT | 7.7 hours | 10.5 hours | 14.5 |
| Transplant # | 1st | 2nd | 2nd |
| Donor age range | 20-30 | 60-70 | 70-80 |
| Recipient sex | M | F | M |
| Recipient age range at transplant | 50-60 | 40-50 | 60-70 |
| Recipient ethnicity | Other | black | indo-asian |
| Cause of ESRF | Unknown | FSGS | diabetes |
| HLA Mismatch | 1-1-0 (2/6) | 2-1-0 (3/6) | 1-2-0 (3/6) |
| Medical context | Hepatitis B;<br>Transplant renal<br>artery stenosis | Infections (warts,<br>urinary tract, thrush,<br>COVID); New onset<br>diabetes | Ischaemic heart<br>disease; Cognitive<br>dysfunction; CMV<br>viraemia;<br>pancytopenia |
| Creatinine at Biopsy<br>($\mu\text{mol/L}$) | 217 | 127 | 250 |
| WBC counts at Biopsy<br>($\times 10^9/\text{L}$) | 5.3 | 12.7 | 1.3 |
| Neutrophils | 3.7 | 9.1 | 0.9 |
| Lymphocytes | 0.6 | 2.0 | 0.3 |
| Monocytes | 0.6 | 1.3 | 0.0 |
| Eosinophils | 0.3 | 0.3 | 0.1 |
| Basophils | 0.0 | 0.0 | 0.0 |
| RBC count at Biopsy | 4.53 | 4.05 | 2.89 |
| (x10<sup>12</sup>/L) |  |  |  |
| PLT count at Biopsy (x10<sup>9</sup>/L) | 263 | 290 | 79 |
| Sensitised (calculated reaction frequency) | No (0%) | Yes (43%) | Yes (0%) |
| Induction agent | basiliximab | alemtuzumab | alemtuzumab |
| Maintenance immunosuppression | FK, MMF | FK | FK, MMF |
| Diagnostic Biopsy data (features in sample examined for diagnosis) |  |  |  |
| Time of biopsy post-transplant | ~ 4 months | ~ 76 months | ~ 4 months |
| anti-HLA DSA @ time of biopsy | Negative | Negative | Negative |
| Composition of Light microscopy sample for diagnosis | 2 cores 16 and 20 mm composed of cortex and medulla | 2 cores 14 and 17 mm composed of cortex and medulla | 2 cores 11 and 12 mm composed of cortex |
| Main diagnosis | Polyoma virus nephropathy | Acute tubular injury | Borderline for T cell-mediated rejection |
| % of cortex involved by tubular atrophy and interstitial fibrosis | 30 | 30 | 10 |
| Immunofluorescence findings, if performed for diagnosis | n/a | Scanty mesangial IgM deposition | n/a |
| Diagnostic EM findings, if performed for diagnosis | n/a | Mesangial EDD; endothelial tubuloreticular inclusions; cg1a | No EDD; cg1a |
| Banff lesion scores* |  |  |  |
| t | 3 | 0 | 2 |
| i | 3 | 0 | 1 |
| v | 0 | 0 | 0 |
| g | 0 | 0 | 0 |
| ptc | 1 | 0 | 0 |
| i-IFTA | 3 | 1 | 2 |
| ti | 3 | 0 | 1 |
| ct | 2 | 2 | 1 |
| ci | 2 | 2 | 1 |
| cv | 0 | 0 | 1 |
| cg | 0 | 1a | 1a |
| ah | 0 | 0 | 1 |
| PTCML | n/a | 3 x 5 layers (mean 2.92 layers) | 0 x 5 layers (mean 2.08 layers) |
| C4d | 0 | 0 | 0 |

Abbreviations: DBD: deceased after brain death; DCD deceased after cardiac death; CIT: cold ischaemic time; M: male; F: female; FSGS: focal and segmental glomerulosclerosis; HLA: human leucocyte antigen; WBC: white blood cell; RBC: red blood cell; PLT: platelet; CRF: calculated reaction frequency; FK: tacrolimus; MMF: mycophenolate mofetil; DSA: donor specific antibody; IF: immunofluorescence; EDD: electron dense deposits; \*t, i, v, g, ptc, i-IFTA, ti, ct, ci, cv, cg, ah, PTCML, C4d are semi-quantitative grading of activity and chronicity (c) according to the Banff Classification for Allograft Pathology; respectively: tubulitis, interstitial inflammation non-scarred cortex, endarteritis, glomerulitis, peritubular capillaritis, interstitial inflammation scarred cortex, interstitial inflammation total cortex; chronic tubular changes, chronic interstitial changes, chronic vascular changes, glomerular capillary wall double contours, arteriolar hyalinosis, peritubular capillary basement membrane multilayering and C4d (complement factor 4d) immunoperoxidase.

Patient 1 was a man in his 50s, hepatitis B carrier, with end stage renal failure (ESRF) due to unknown cause. He developed a post-transplant renal artery stenosis that was stented, then a rise in creatinine triggered biopsy 1 at 136 days post-transplant which showed extensive tubulointerstitial inflammation related to viral infection of the graft by polyoma virus with chronic tubulointerstitial fibrosis amounting to about 30% of cortex. This was treated by stopping the patient’s MMF, with subsequent improvement in renal function.

Patient 2 was a woman in her 50s with ESRF attributed to familial FSGS. She had a first transplant almost 20 years previously, which eventually failed, followed by a second transplant. After the second transplant she suffered from many immunosuppression-related infections (warts, urinary tract, thrush, COVID) and new onset diabetes. A slow upward drift in her creatinine levels around 6.5 years post-transplant led to biopsy 2, which showed acute tubular injury as the main diagnostic active feature, with chronic tubulointerstitial fibrosis amounting to about 30% of cortex. Her creatinine stabilised but did not improve.

Patient 3 was a man in his 60s who had ESRF due to diabetic nephropathy. He also suffered from cognitive dysfunction and ischaemic heart disease. He had also had a previous graft, which failed due to recurrent diabetic nephropathy. The current second graft was complicated post-operatively by CMV viraemia and pancytopaenia attributed to immunosuppressive and infection-prevention drugs as he recovered good levels with administration of GM-CSF. A rise in creatinine triggered biopsy 3 at 130 days post-transplant, which showed active inflammation that was ‘borderline’ (i.e. uncertain) for T cell-mediated rejection according to the Banff Classification for Allograft Pathology (Naesens et al., 2024), and chronic tubulointerstitial fibrosis amounting to about 10% of cortex. This was treated with the addition of MMF and steroids to his baseline tacrolimus, and creatinine decreased though not to baseline levels.

### Whole biopsy imaging using High Throughput Tomography (HiTT)

Imaging the nanoscale features of a whole biopsy using volume EM would take decades. To reduce this to a reasonable and practical timeframe, the nanopathology pipeline employs a new synchrotron-based hard X-ray imaging modality called HiTT (High Throughput Tomography) (Albers et al., 2024), implemented on the EMBL P14 beamline at the PETRA III synchrotron on the DESY campus in Hamburg, Germany (Fig.2). HiTT enables biopsies to be imaged intact with sub-micron voxel size. HiTT uses phase contrast, so biopsy tissue can be imaged fully hydrated in fixative, without the need for heavy metal stains, thus preserving antigenicity for downstream molecular labelling. In addition, HiTT is fast, with each field of view taking less than 3 minutes for data acquisition and reconstruction. HiTT therefore provides a high throughput method for imaging the microstructural features of immune-mediated transplant rejection.

**Figure 2.**
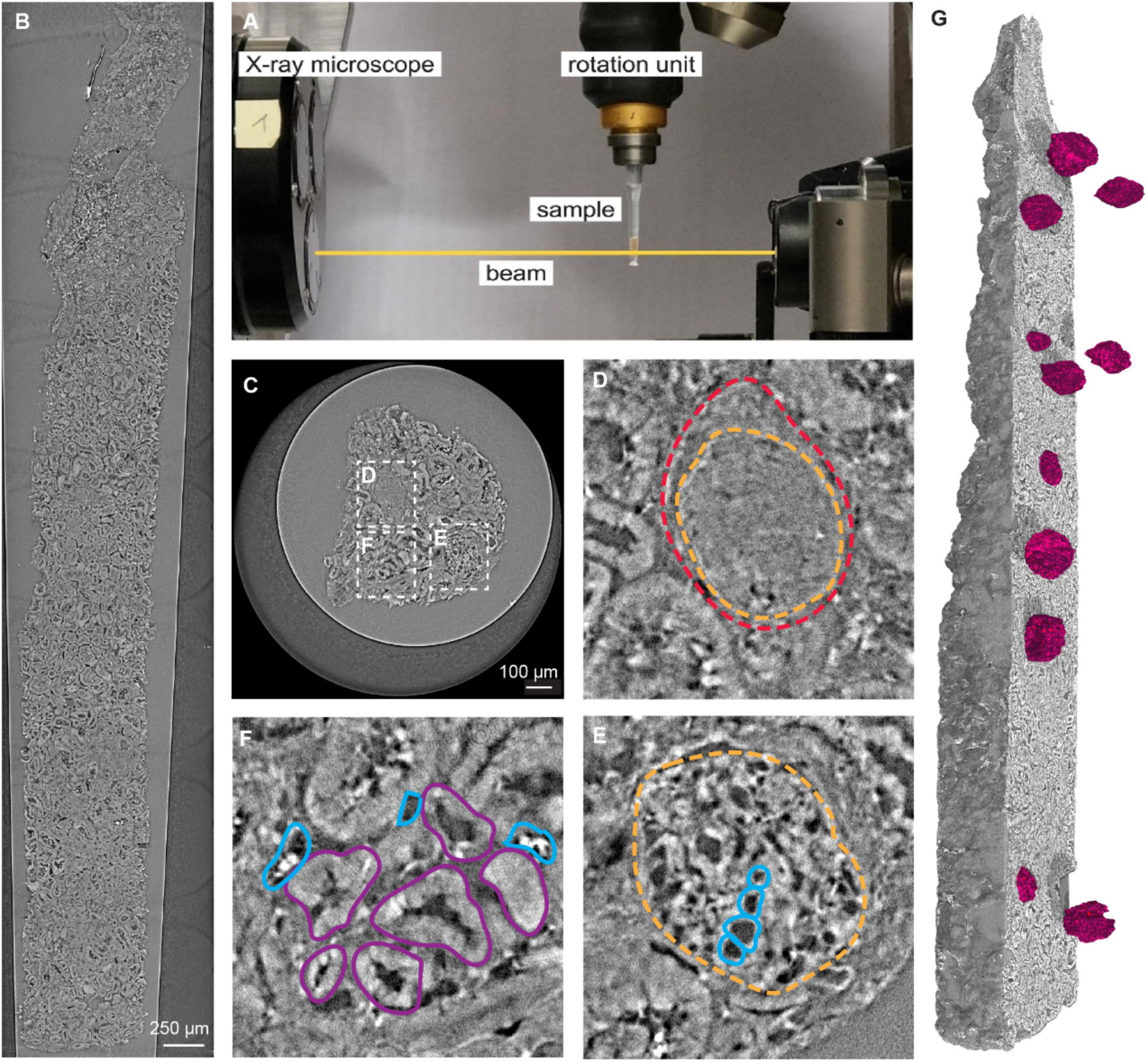
Whole biopsy imaging using High Throughput Tomography (HiTT) (patient 2). A) Side view of the P14 beamline setup for HiTT. The kidney biopsy was mounted in 1% FA in 0.1M PB in a sterile p10 pipette tip and imaged over a 180° rotation in the path of the X-ray beam. B) Longitudinal orthoslice through the reconstructed tomogram, stitched together from a tiled acquisition of 7 individual HiTT scans. C) Transverse orthoslice through the reconstructed tomogram. D) Globally sclerosed glomerulus, with glomerular tuft outlined by the orange dashed line, and Bowman’s capsule outlined by the red dashed line. E) ‘Open’ glomerulus, with the glomerulus outlined by the orange dashed line, and examples of open capillary loops outlined in blue. F) Peritubular capillaries, with example PTCs outlined in blue (some containing red blood cell appearing as bright white objects) and example tubules outlined in purple. G) Volume rendering of the tomogram, with the interior exposed using a clipping plane, and manually segmented glomeruli shown in magenta.

The three selected patient biopsies were photographed and cut (if required) to reduce the tissue length to <6 mm for stability during HiTT imaging. Biopsies were transported to the synchrotron in temperature-controlled shippers to prevent freeze-thaw damage to tissue ultrastructure. The biopsies were loaded into pipette tips and imaged in fixative using a tiled acquisition (Fig.2A). After reconstruction of the data (Fig.2B-D), tomograms were converted into movie files (Supp. Movies 1-3) to support easier viewing of large datasets to screen for microscale features of interest.

Biopsy volume and surface area were calculated. Glomerular segmentation, counts and classification were performed (Fig.2E-G; Table 2). Biopsies from patients 1,2 and 3 contained 23, 14 and 32 glomeruli in total, which could be roughly classified into open, segmentally sclerosed and globally sclerosed capillary loops (Table 2 and Supp.Fig.2). The number of glomeruli was higher than expected for a ∼6 mm length of cortex from experience in diagnostic practice. This is likely because the entire biopsy volume was analysed, contrary to diagnostic pathways that only examine a few percent of the biopsy volume (usually a set of up to ten 1-micron thick slices). In addition to the larger anatomical features that could be visualised with HiTT (glomeruli, convoluted tubules, arteries) it was also possible to see finer microstructural details including peritubular capillaries, red blood cells and the nuclei of some cells.

**Table 2.**
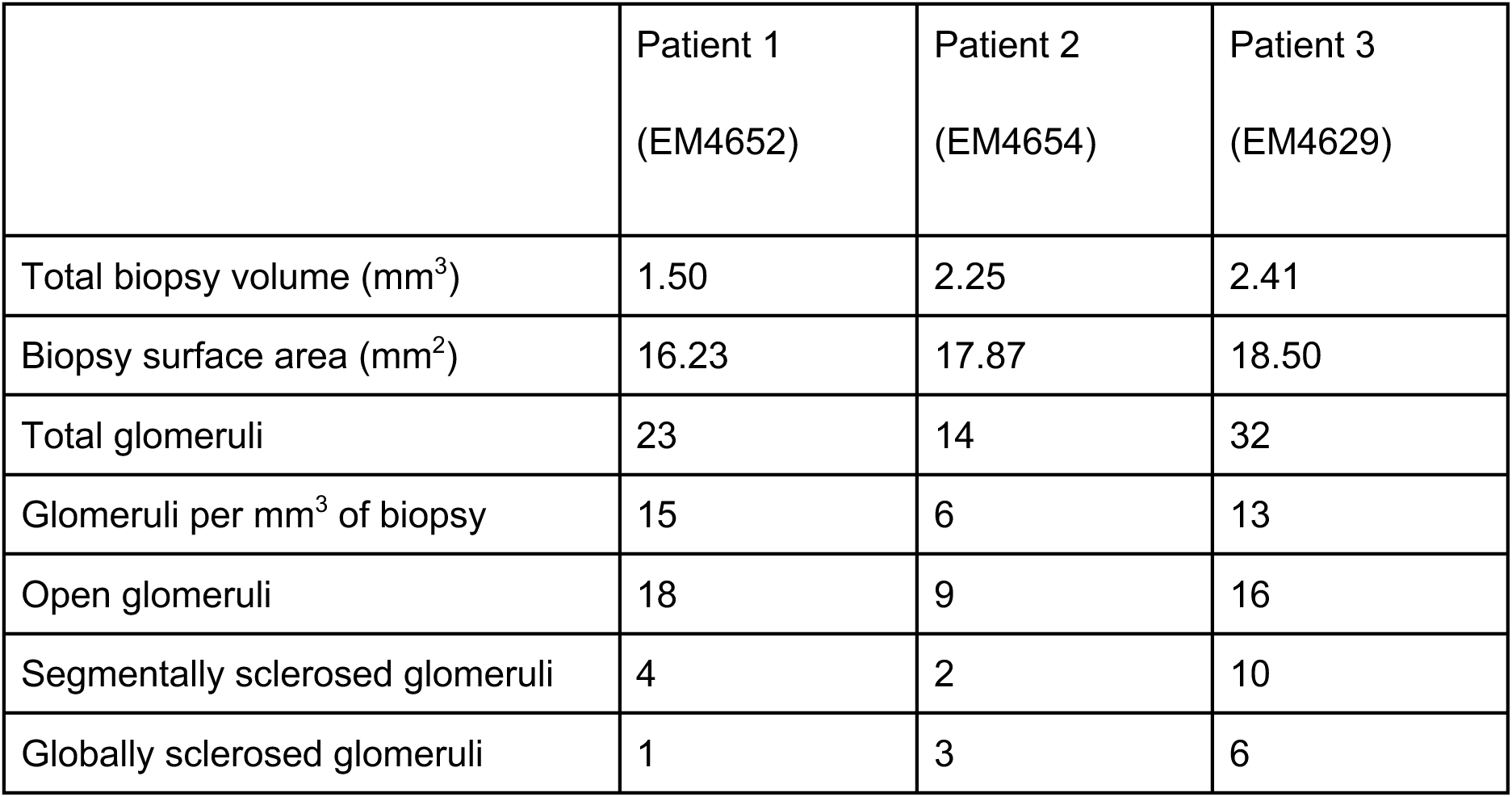
HiTT biopsy metrics.

|  | Patient 1<br>(EM4652) | Patient 2<br>(EM4654) | Patient 3<br>(EM4629) |
| --- | --- | --- | --- |
| Total biopsy volume (mm<sup>3</sup>) | 1.50 | 2.25 | 2.41 |
| Biopsy surface area (mm<sup>2</sup>) | 16.23 | 17.87 | 18.50 |
| Total glomeruli | 23 | 14 | 32 |
| Glomeruli per mm<sup>3</sup> of biopsy | 15 | 6 | 13 |
| Open glomeruli | 18 | 9 | 16 |
| Segmentally sclerosed glomeruli | 4 | 2 | 10 |
| Globally sclerosed glomeruli | 1 | 3 | 6 |

Though the number of patients and biopsies analysed was too low to extract any statistically significant quantification of biopsy and glomerular features, the proof of principle that these measures can be extracted from HiTT data demonstrates the potential of the technique for larger scale clinical morphometric studies. In terms of targeting ROIs, the HiTT data enabled precise selection of glomerular and peritubular capillary coordinates for downstream molecular and ultrastructural analysis. HiTT also provided microscale landmarks for registration of correlative multimodal image datasets.

### Targeting strategy for molecular imaging

Movies of the HiTT volumes were used to select a suitable glomerulus and adjacent peritubular capillaries (without obvious sclerosis/ fibrosis) in each biopsy (Fig.3 and Supp.Fig.3). To find and unequivocally identify immune cells within these structures, immunolabelling for immune cell surface antigens was required. Immunofluorescence labelling was preferred over immunogold labelling, since fluorescence microscopy supports screening over larger areas than electron microscopy, and antibodies compatible with EM immunolabelling are rare. However, immunofluorescence labelling usually requires permeabilisation agents (e.g. methanol, Triton X-100) to allow penetration of antibody conjugates into the sample, with the consequence that ultrastructure is compromised. To avoid compromised ultrastructure, the biopsies were embedded in low melting point agarose and sliced at room temperature using a vibrating blade microtome (vibratome). A slice thickness of 60 µm was found to be suitable for both reagent penetration and imaging in the fluorescence microscope.

**Figure 3.**
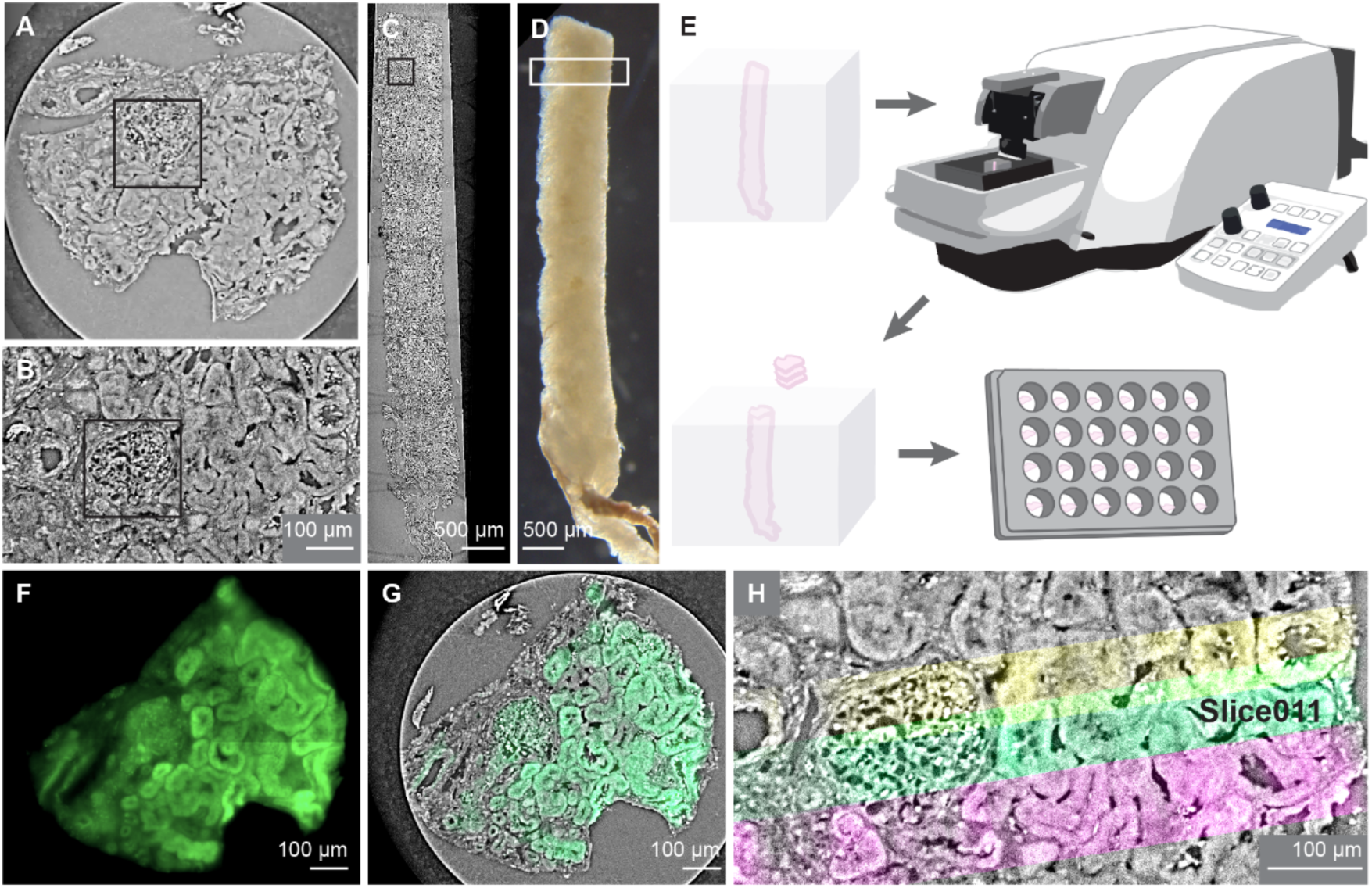
Targeting strategy for molecular and ultrastructural imaging (patient 2). A) Transverse orthoslice through the reconstructed HiTT tomogram showing a glomerulus (box) selected for downstream molecular and ultrastructural imaging. B) Selected glomerulus (box) displayed in the longitudinal plane of the biopsy and C) zoomed out to show the selected glomerulus in the context of the whole biopsy. D) Photograph of the biopsy showing an approximate location of the selected glomerulus (white box) in comparison to the hydrated fixed HiTT data from the same biopsy shown in panel C. E) The biopsy was embedded vertically in a cube of low melting point agarose with the selected glomerulus at the top. The sample was mounted in the vibratome with the glomerulus facing the blade and sequential 60 μm thick slices were cut and transferred in sequence to a 24-well plate. F) Widefield autofluorescence image of vibratome slice 11 containing a large part of the selected glomerulus. G) Overlay of the widefield autofluorescence data (green) onto the fixed hydrated biopsy HiTT data (greyscale) for vibratome slice 11. H) Widefield autofluorescence data for slices 10 (yellow), 11 (green) and 12 (pink) containing the selected glomerulus, aligned to the fixed hydrated biopsy HiTT data (greyscale), shown in the longitudinal plane.

For targeted vibratome sectioning, the approximate position of the selected ROI (Fig.3A,B and Supp.Fig.3A,B,F,G) was first identified in the intact biopsy by comparing the morphology of the biopsy as seen in the HiTT data (Fig.3C and Supp.Fig.3C,H) to that seen using a dissection microscope (Fig.3D). The biopsy was then mounted so that the selected ROI faced the knife in the vibratome, and the approximate number of vibratome slices needed to reach and slice through the ROI was calculated. Sections were then cut, collected and stored in order in a 24-well plate (Fig.3E). Tissue autofluorescence was imaged (Fig.3F and Supp.Fig.3D,I) and compared to the HiTT data (Fig.3G and Supp.Fig.3E,J) to confirm the content of each vibratome slice. Autofluorescence images of sequential vibratome slices into the HiTT volume demonstrated continuity of slices in the longitudinal plane (Fig.3H).

### Molecular imaging using non-permeabilisation immunofluorescence

Vibratome slicing effectively opened blood vessels and tubules so that antibodies and other reagents could reach the surface of immune cells in the vessel lumina without the need for permeabilisation agents (Fig.4 and Supp.Figs 4 & 5).

**Figure 4.**
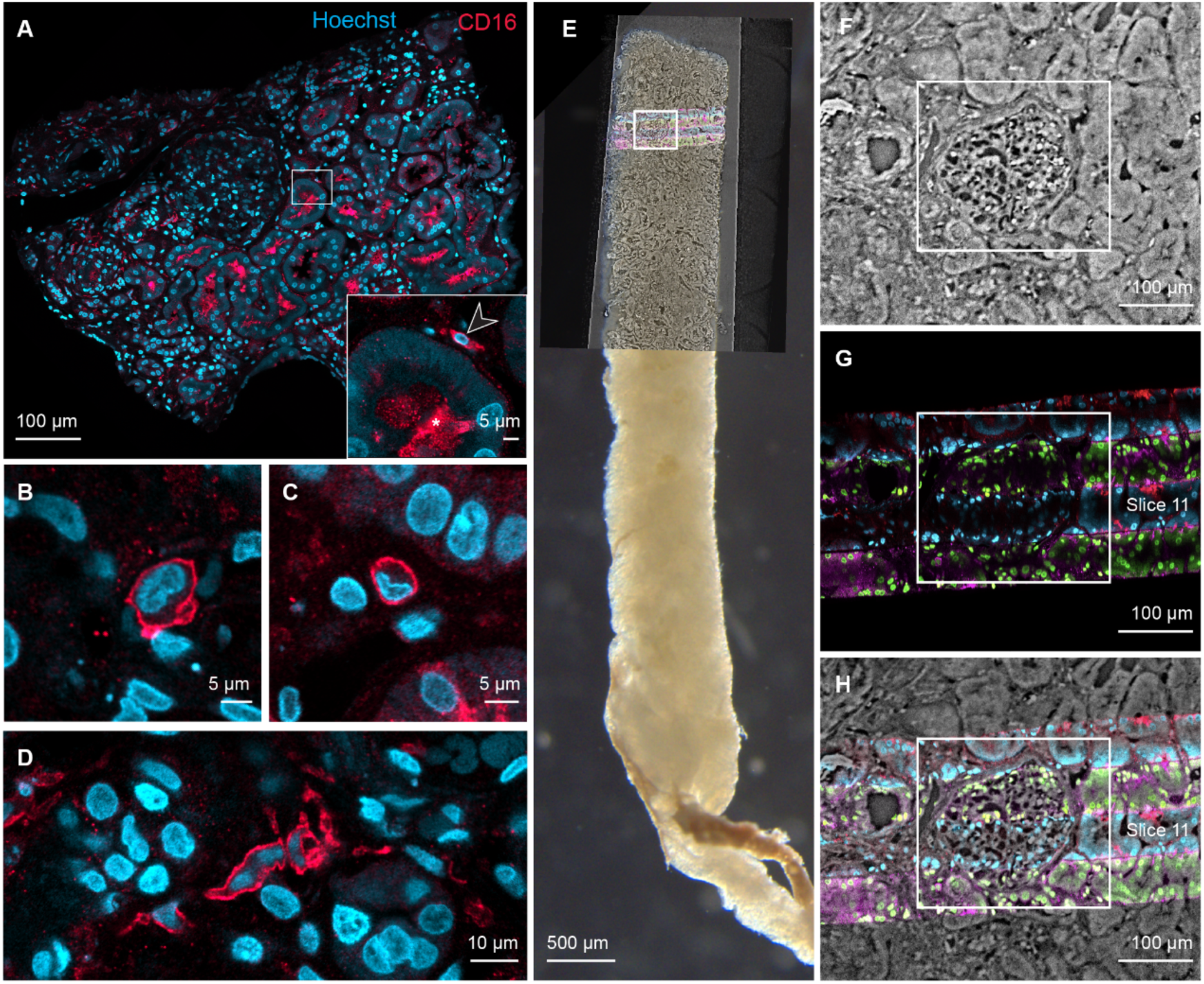
Molecular imaging using non-permeabilisation immunofluorescence (patient 2). A) Confocal fluorescence image of vibratome slice 11 showing CD16-AF647 (red) and Hoechst H33342 (blue). Inset: Non-specific fluorescence signal within the lumen of tubules (asterisk) could be distinguished from CD16-positive immune cells (arrowhead) based on location and morphology. B) CD16-positive immune cell within the glomerulus in slice 11. C) CD16-positive immune cell within a peritubular capillary in slice 11. D) CD16-positive immune cell within the interstitium in slice 11. E) Confocal fluorescence data from four serial vibratome slices (slices 9-12) registered to the fixed hydrated biopsy HiTT data and the photograph of the biopsy (confocal slices are shown perpendicular to the original imaging plane). The target glomerulus is highlighted (box). To clearly differentiate the four slices when registered into the volume, different colours were applied to the images. Slices 9 and 11 are displayed with Hoechst H33342 signal in blue and CD16 signal in red; slices 10 and 12 are displayed with Hoechst H33342 signal displayed in green and CD16 signal displayed in magenta. F-H) The target glomerulus identified in the HiTT data (F; box) is contained within the four vibratome slices imaged by confocal fluorescence microscopy (G) with minimal loss of material between adjacent slices highlighted by overlaying the fluorescence onto the HiTT data (H).

**Figure 5.**
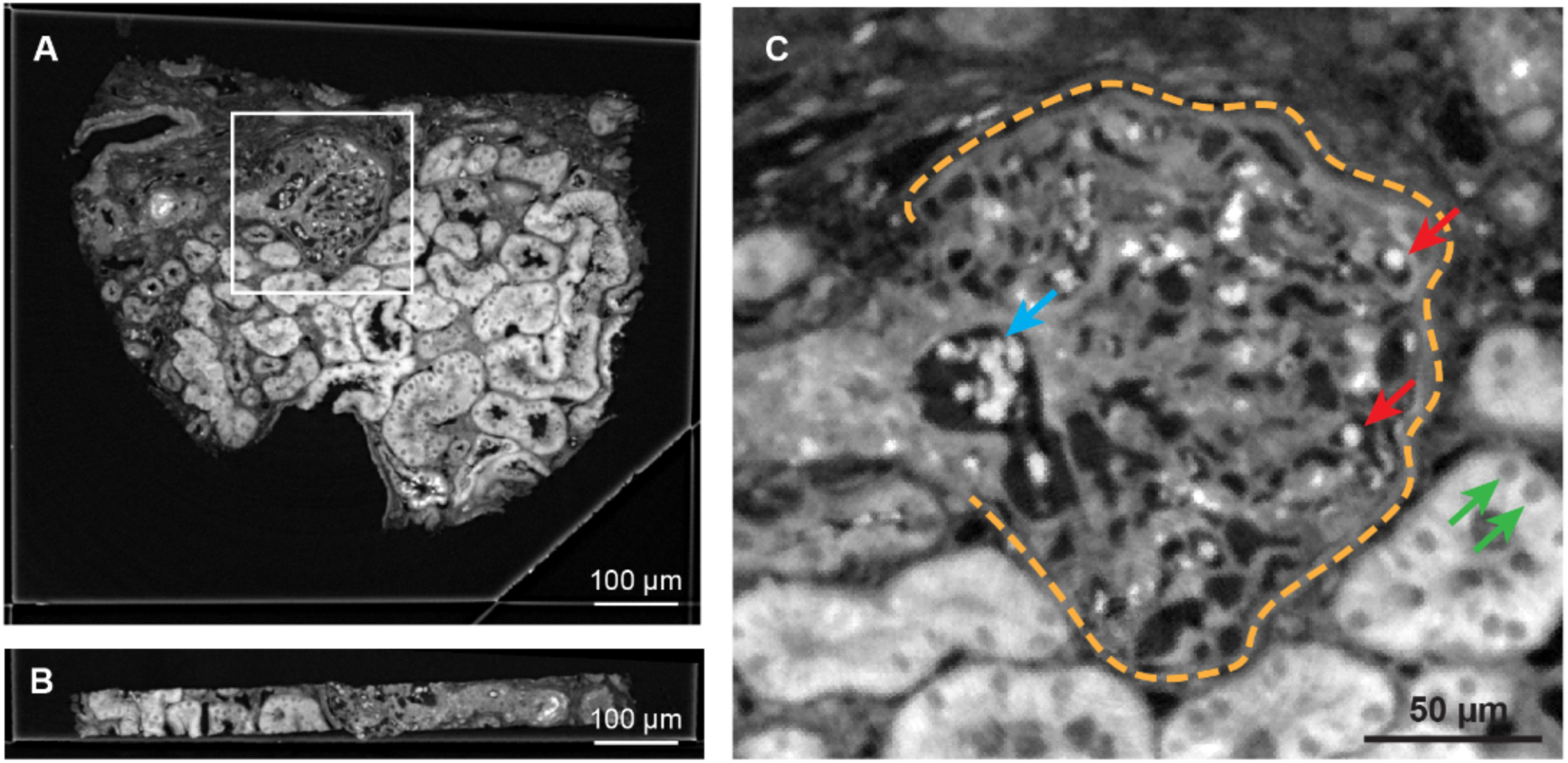
Post-embedding HiTT of heavy metal stained, resin embedded vibratome slices (patient 2). A) An ‘*en face*’ 2D plane from the 3D tomogram of the vibratome slice showing key structures including a glomerulus and peritubular capillaries. B) A side view of the same 60 µm thick slice. C) The increased contrast in the samples delivered improved contrast and resolution in the HiTT datasets. The glomerulus is outlined with an orange dashed line. Red blood cells in the glomerular capillary loops (red arrows) and hilar capillary (blue arrow), and nuclei of the tubular epithelium cells (green arrows) are clearly visible.

Vibratome slices from the selected ROI from each of the patient biopsies were immunolabelled and imaged using confocal fluorescence microscopy (Fig.4A, Supp.Fig.4A, Supp.Fig.5A), revealing the location of CD16-positive immune cells within glomeruli (Fig.4B, Supp.Fig.4C, Supp.Fig.5C), peritubular capillaries (Fig.4C, Supp.Fig.4B, Supp.Fig.5B), and interstitium (Fig.4D). CD16 labelled the plasma membrane of the immune cells only, and not intracellular pools of CD16, because no permeabilisation agent was used. The membrane permeant dye Hoechst was also added to label cell nuclei, to be used downstream as landmarks for registration of correlative multimodal datasets (Fig.4A-D, Supp.Fig.4A-C, Supp.Fig.5A-C).

Fluorescence images from serial vibratome slices were registered to the fixed hydrated HiTT data (Fig.4E, Supp.Fig.4D, Supp.Fig.5D), confirming that little (if any) tissue was lost between slices (Fig.4F-H, Supp.Fig.4E-G, Supp.Fig.5E-G).

### Targeted volume EM of ROIs

The resin embedded slices were imaged using X-ray microscopy (XRM) to relocate the ROIs within the tissue and extract coordinates for targeted volume EM using SBF-SEM (Fig.5 and Supp.Fig.6). Due to heavy metal staining and resin embedding, the samples had more contrast than the fixed hydrated biopsies, so could be imaged using either laboratory-based microCT or synchrotron-based HiTT, though HiTT was faster. Features including red blood cells in the vessel lumina and cell nuclei in the tubules were clearly visible.

**Figure 6.**
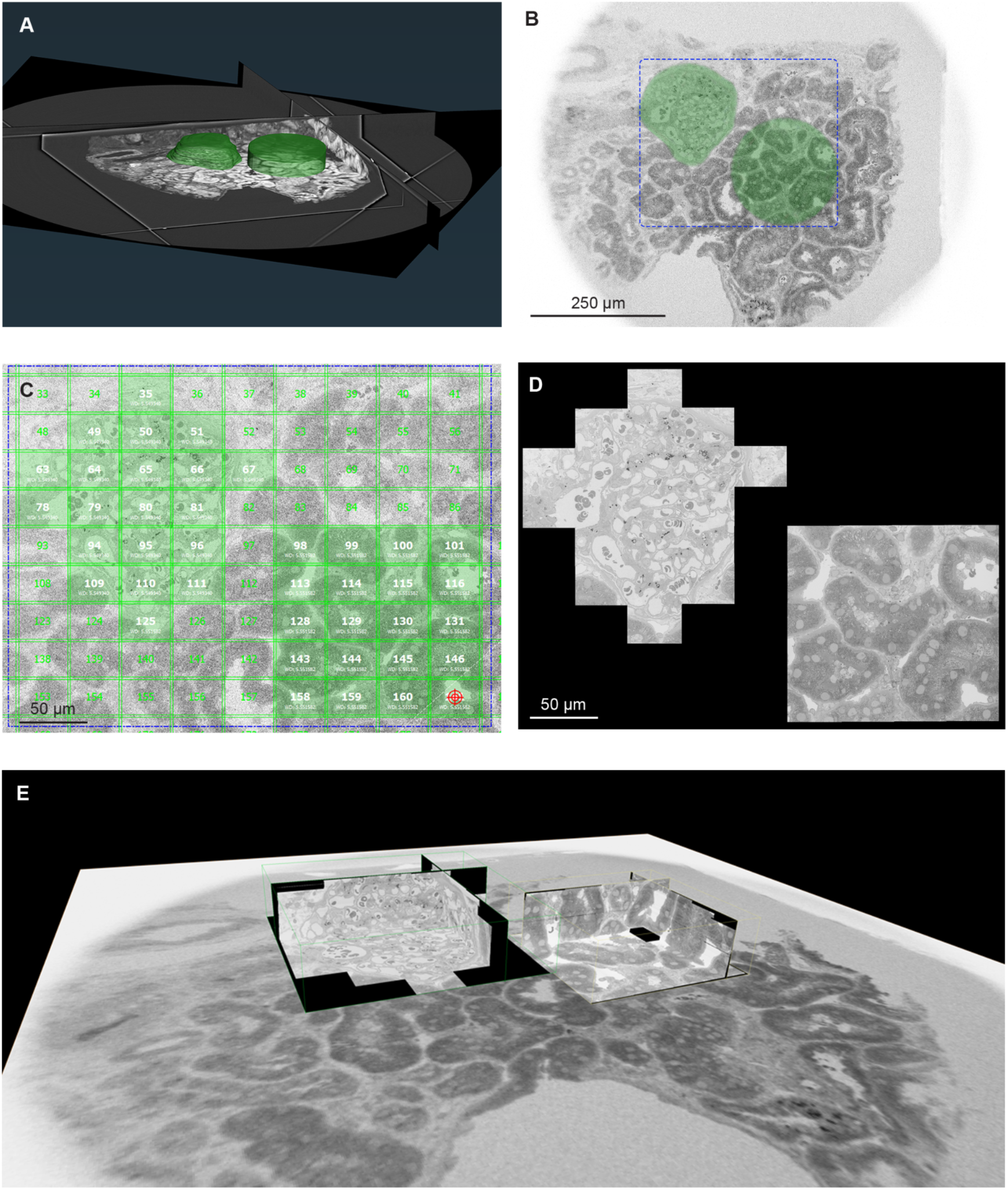
Resin-HiTT-targeted SBF-SEM imaging of a glomerulus ROI and a PTC-rich ROI (patient 2). A) 3D visualization of the HiTT data acquired from the heavy metal stained, resin embedded vibratome slice. Two regions of interest (ROIs) containing a glomerulus and a region rich in peritubular capillaries were identified and segmented (green) in the HiTT data. B) Low resolution overview image of the block surface acquired using SBF-SEM. The two ROIs identified in the HiTT volume are shown (green masks). The dashed blue box highlights the area shown in panels C and D. C) Screenshot of the SBEMimage interface showing a grid that covers the entire imaging field. Active tiles (green) differ for each block surface exposed during cutting according to the size and position of the ROIs. The crosshair (red) shows the current stage position. D) Post-acquisition stitching and alignment of the tiles produces large high resolution images of the ROIs. E) High resolution volumes of the 2 ROIs registered and overlaid with low resolution overviews to show their position in context.

For each slice, the ROIs (matching volumes of glomerulus and peritubular capillaries) were manually segmented in the XRM data (Fig.5A, Supp.Fig.7A, Supp.Fig.8A) to provide coordinates for imaging in the SBF-SEM (Fig.5B, Supp.Fig.7B, Supp.Fig.8B). The open-source software SBEMimage (Titze et al., 2018) was used to control SBF-SEM image acquisition from the selected ROIs (Fig.5C-E, Supp.Fig.7C,D, Supp.Fig.8C,D). Images from XRM and SBF-SEM had similar contrast and could be easily registered to drive automated switching between imaging parameters in the fast lower resolution approach phase and slower high resolution ROI imaging phase in the SBF-SEM. SBF-SEM imaging was monitored on-the-fly by comparing the block surface image to the XRM data.

**Figure 7.**
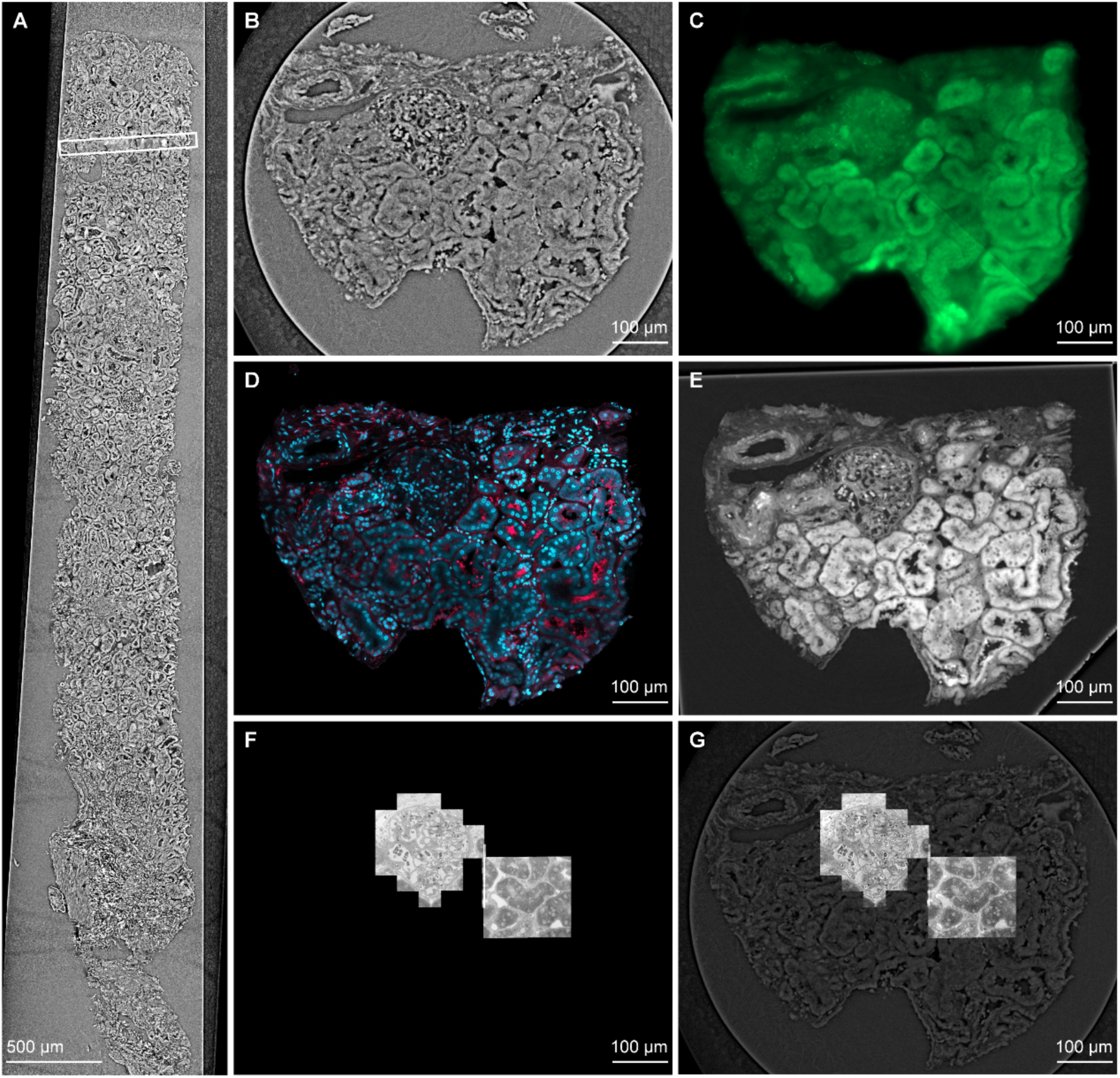
Multimodal imaging of a human kidney biopsy using the nanopathology pipeline (patient 2). A) Longitudinal image of a fixed hydrated biopsy taken from the tomogram acquired using HiTT. The box highlights the vibratome slice that was processed through the entire nanopathology pipeline. B) Transverse image taken from the tomogram acquired using HiTT in the region of the vibratome slice that was processed through the entire nanopathology pipeline. C) Autofluorescence image of the fixed hydrated vibratome slice. D) Confocal fluorescence image of the fixed hydrated vibratome slice labelled with CD16-AF647 (red) and Hoechst H33342 (blue). E) HiTT of the resin embedded vibratome slice. F) SBF-SEM of the ROIs from the resin-embedded vibratome slice. G) SBF-SEM data overlaid onto the fixed hydrated HiTT data to add tissue context.

**Figure 8.**
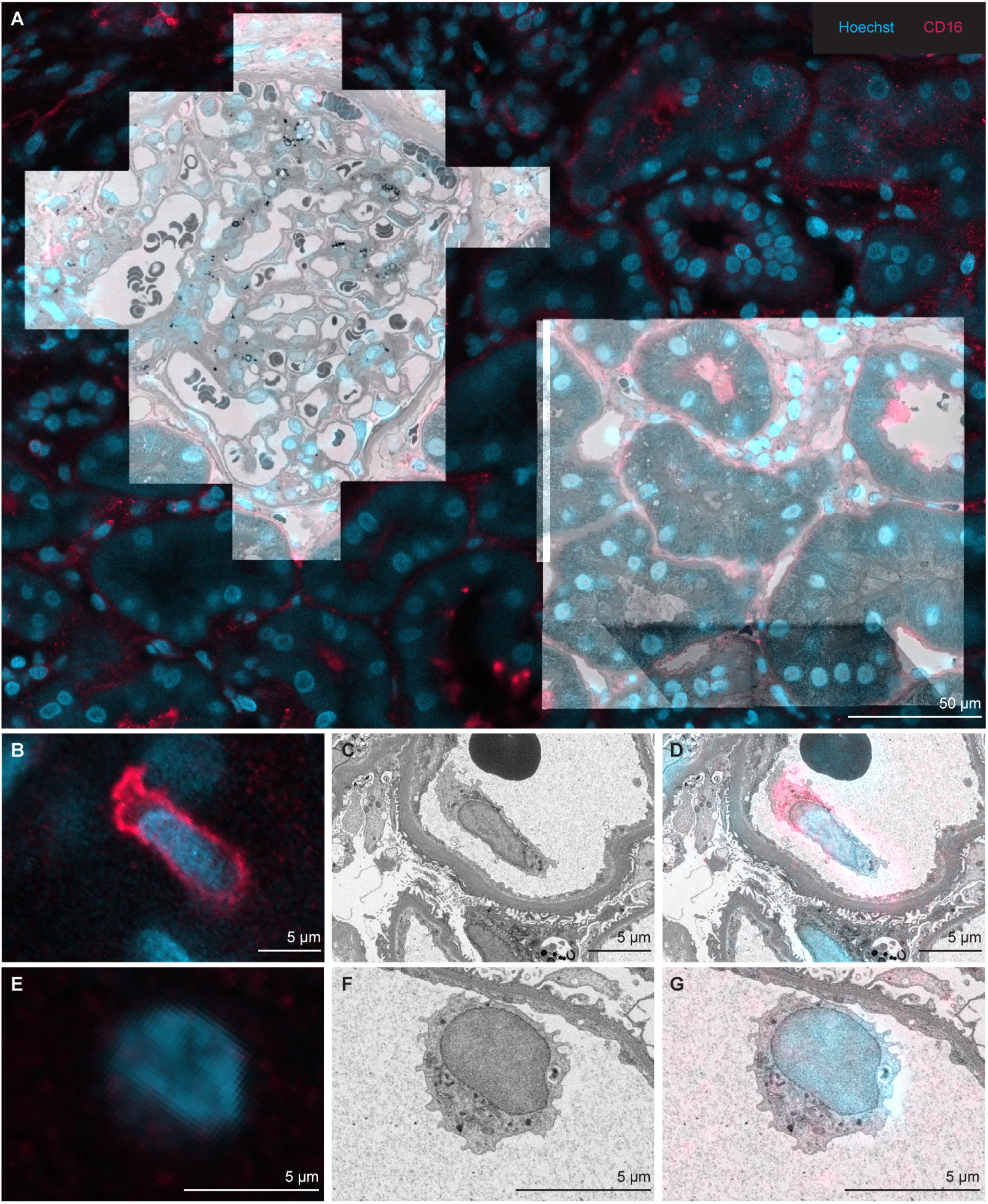
Locating and identifying immune cells via correlation of multimodal data in MoBIE (patient 2). A) 2D snapshot of confocal fluorescence (CD16-AF647 in red and Hoechst H33342 in blue), resin HiTT and SBF-SEM data registered in MoBIE using cell nuclei as landmarks. B-D) CD16-positive immune cell in fluorescence microscopy (panel B), SBF-SEM (panel C) and overlaid fluorescence and EM data (panel D). E-G) CD16-negative immune cell in fluorescence microscopy (panel E), SBF-SEM (panel F) and overlaid fluorescence and EM data (panel G).

The tiles acquired using SBF-SEM were stitched to create image volumes representing the glomerular and PTC ROIs. The glomeruli and PTC ROIs were segmented manually and meshes created to allow downstream calculation of the number of immune cells per volume of glomerulus/ PTC imaged for each slice.

### Correlation of multimodal data in MoBIE

The nanopathology pipeline generated at least five image datasets per biopsy (Fig.7): Fixed hydrated biopsy HiTT (Fig.7A,B), autofluorescence of vibratome slices (Fig.7C), confocal fluorescence of vibratome slices (Fig.7D), microCT or HiTT of resin embedded vibratome slices (Fig.7E), and SBF-SEM of vibratome slices (Fig.7F,G).

The Fiji plugin MoBIE (Pape et al., 2023) was used to register and view all image datasets acquired from the same sample in the same virtual space (Fig.8, Supp.Figs 9 & 10). MoBIE supported remote navigation of the registered multimodal datasets so that end users could seamlessly explore terabyte-scale image data to find and analyse immune cells in the context of the whole biopsy structure at the milli-, micro-and nano-scales (Fig.8A, Supp.Fig.9A, Supp.Fig.10A). Immune cells could be located and unequivocally assigned as CD16-positive monocytes (Fig.8B-D, Supp.Fig.9B-D, Supp.Fig.10B-D) or CD16-negative immune cells (Fig.8E-G, Supp.Fig.9E-G, Supp.fig.10B-D).

**Figure 9.**
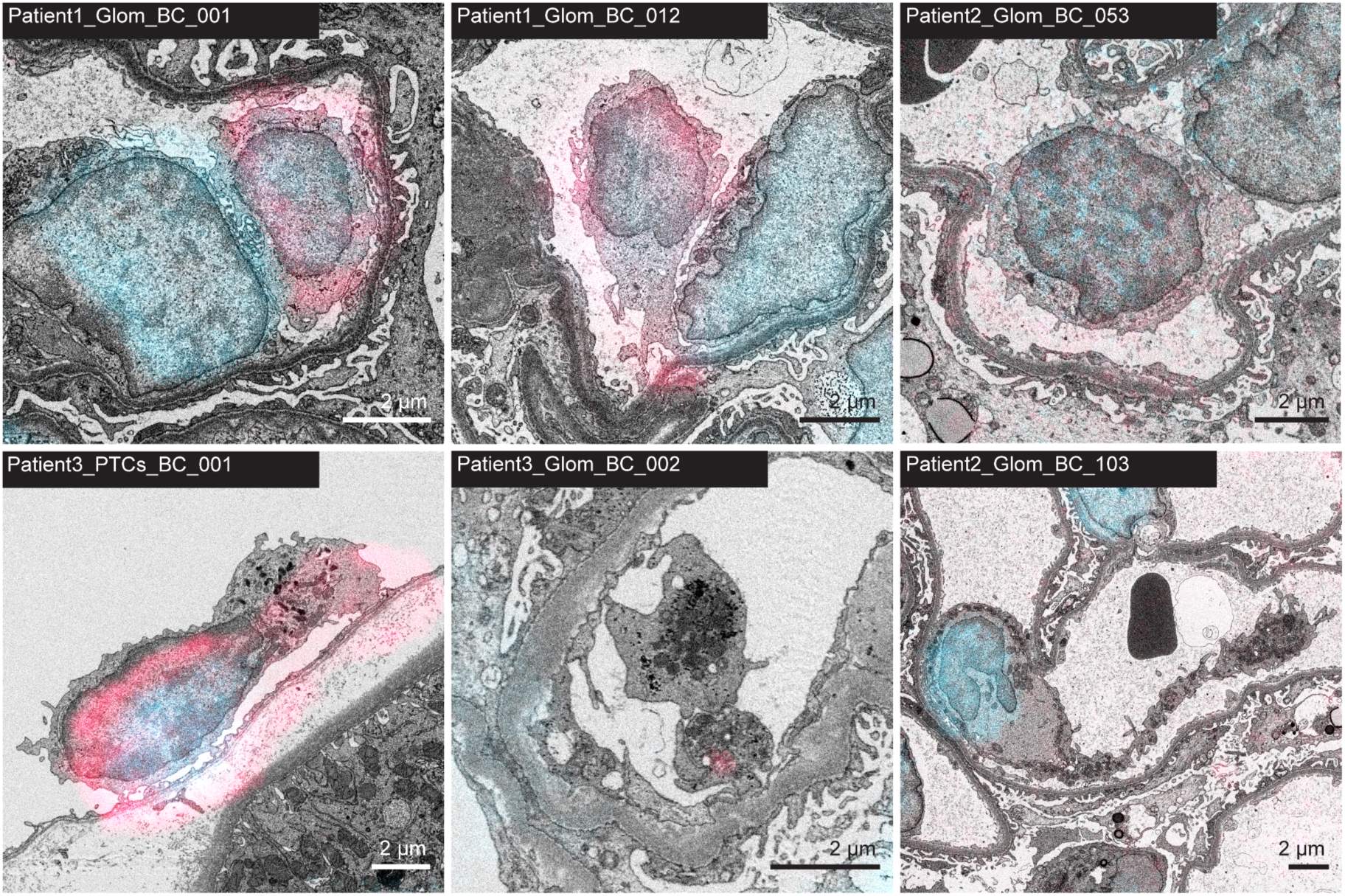
Gallery of immune cells located and imaged using the nanopathology pipeline. First row, from left to right: patient 1, CD16-positive mononuclear cell establishing a synapse with an adjacent endothelial cell in a glomerular capillary (Supp. Movie 4); patient 1, CD16-positive mononuclear cell patrolling the endothelial surface and subendothelial space in a glomerular capillary (Supp. Movie 5); patient 2, CD16-negative mononuclear cell patrolling the endothelial surface in a glomerular capillary (Supp. Movie 6). Second row, from left to right: patient 3, CD16-positive mononuclear cell patrolling the endothelial cell surface in a peritubular capillary (Supp. Movie 8); patient 3, intraluminal platelets with cell surface projections establishing contacts with each other and with the adjacent endothelium in a glomerular capillary (Supp. Movie 9); patient 2, megakaryocyte with a long tail of budding platelets in a glomerular capillary (Supp. Movie 7).

Correlation across multimodal datasets revealed that immune cells identified using SBF-SEM were also visible in the resin HiTT and to some extent in the fixed hydrated HiTT images (Supp.Fig.11). Immune cells were clearly visible in HiTT when located within the vessel lumen but were less clear when in close proximity to the endothelium or when located in the interstitium. Immune cells were easier to locate in resin HiTT than in fixed hydrated HiTT due to the increased contrast and resolution of the former.

### Quantification of immune cells in patient transplant biopsies

Individual blood cells were manually quantified and classified per volume of glomerulus examined and in the peritubular capillary network (Table 3, Supp. Table 1, Supp. Figs.12-14). The total number of circulating cells (not counting red blood cells) per volume unit was similar for glomeruli (range = 21.4 - 135.7 x 10^-6^ /µm^3^) and peritubular capillaries (range = 27.7 - 167.6 x 10^-6^ /µm^3^). The predominant blood cell type in glomeruli was platelets in all three patients (range = 8.8 - 103.5 x 10^-6^/µm^3^), followed by small mononuclear cells in all three patients (range = 7.6 - 13.3 x 10^-6^/µm^3^). This was followed by large mononuclear cells with abundant cytoplasm, then neutrophil polymorphs in patients 1 and 3, whereas patient 2 had more neutrophil polymorphs than large mononuclear cells. Volume examined and therefore cell numbers were overall smaller in peritubular capillaries, and only small mononuclear cells and platelets were observed.

**Table 3.** Manual Quantification of Intraluminal cell in ROI.

|  | Patient 1<br>(EM 4652) | Patient 2<br>(EM 4654) | Patient 3<br>(EM 4629) |
| --- | --- | --- | --- |
| **Glomerulus** |  |  |  |
| Volume of glomerulus examined with SBF-SEM | 793,459 (793,500) $\mu\text{m}^3$<br>(of 3,340,040 $\mu\text{m}^3$ whole glom volume) | 831,196 (831,200) $\mu\text{m}^3$<br>(of 2,599,920 $\mu\text{m}^3$ whole glom volume) | 375,935 (375,900) $\mu\text{m}^3$<br>(of 3,137,200 $\mu\text{m}^3$ whole glom volume) |
| Mononuclear cells, small<br>[per $10^{-6}/\mu\text{m}^3$] | 6 (5 CD16-pos, 1 CD16-neg)<br>[7.6] | 11 (6 CD16-pos, 5 CD16-neg)<br>[13.2] | 5 (4 CD16-pos, 1 CD16-neg)<br>[13.3] |
| Mononuclear cells, large<br>[per $10^{-6}/\mu\text{m}^3$] | 3 (1 CD16-pos, 2 CD16-neg)<br>[3.8] | 5 (3 CD16-pos, 2 CD16-neg)<br>[6.0] | 2 (2 CD16-pos, 1 CD16-neg)<br>[5.3] |
| Polymorphs<br>[per $10^{-6}/\mu\text{m}^3$] | 1<br>[1.3] | 9 (7 CD16-pos, 2 CD16-neg)<br>[10.8] | 1 (CD16-pos)<br>[2.7] |
| PLT<br>[per $10^{-6}/\mu\text{m}^3$] | 7<br>[8.8] | 86<br>[103.5] | 17<br>[45.2] |
| **Total in volume examined with SBF-SEM** | 17 | 112 (1 unclassifiable) | 25 |
| **Total per $10^{-6}/\mu\text{m}^3$** | 21.4 | 134.7 | 66.5 |
| **Total per glomerulus (estimated)** | 42 leukocytes<br>29 PLT | 78 leukocytes<br>269 PLT | 66 leukocytes<br>141 PLT |
| **Peritubular capillaries** |  |  |  |
| Volume of PTCs examined | 65,643 (65,600) $\mu\text{m}^3$ | 60,921 (60,900) $\mu\text{m}^3$ | 72,112 (72,100) $\mu\text{m}^3$ |
| Mononuclear cells, small<br>[per $10^{-6}/\mu\text{m}^3$] | 7 (4 CD16-pos, 3 CD16-neg)<br>[106.6] | 0 | 2 (1 CD16-pos, 1 CD16-neg)<br>[27.7] |
| Mononuclear cells, large<br>[per $10^{-6}/\mu\text{m}^3$] | 0 | 0 | 0 |
| Polymorphs | 0 | 0 | 0 |
| PLT<br>[per $10^{-6}/\mu\text{m}^3$] | 4<br>[60.9] | 6<br>98.5] | 0 |
| **Total in volume examined with SBF-SEM** | 11 | 6 | 2 |
| **Total per $10^{-6}/\mu\text{m}^3$** | 167.6 | 98.5 | 27.7 |

Compared to normal and patient-measured circulating cell counts in peripheral blood, platelets would be expected to predominate, as observed. All three patients had more circulating neutrophils than monocytes and lymphocytes combined, which contrasts with our findings that there were more mononuclear cells than neutrophils seen in the glomeruli. This suggests relative enrichment of mononuclear cells compared to neutrophils in the glomerular capillary loops. Histological assessment of the matched biopsy samples used for diagnosis, according to semi-quantitative Banff lesion scores (as per standard diagnostic practice), suggested that none of these patients had abnormal glomerular inflammation (g0); this is reflected in similar amounts of mononuclear cells overall across all three patients in glomeruli. Patient 1 was recorded as having a mild increase of cells in peritubular capillaries (ptc1), reflected in an overall higher mononuclear cell count.

### 3D nanoscale features of immune cells in patient transplant biopsies

Beyond cell counts, we sought to document cell behaviours previously only described in animal or *in vitro* models of disease or inferred computationally from human samples. Although described here qualitatively, future efforts to segment, classify and quantify cellular and subcellular features will support development of disease models.

### Endothelial cell surface patrolling by CD16-positive monocytes

Intravital microscopy has documented ‘patrolling’ of the kidney endothelial cell surface by CD16-positive monocytes in an LFA-1 dependent manner (Finsterbusch et al., 2016). These innate immune cells carry out immune surveillance, sampling the endothelial surface and subendothelial space for antibodies and immune complexes; in animal models of antibody-mediated injury, CD16-positive monocyte activation is an early feature of disease that orchestrates recruitment of other immune cells and glomerular injury (Turner-Stokes et al., 2020). Single cell RNA sequencing complemented with intercellular communication analysis has found evidence of communication between CD16-positive monocytes and endothelial cells in human transplant biopsies (Lamarthée et al., 2023), but imaging of this behaviour in human samples has not been previously achieved. Here, we document the routine occurrence, in three transplant patients without established rejection, of patrolling of both glomerular and peritubular capillary endothelium by CD16-positive mononuclear cells extending numerous fine cell surface projections to establish contacts with the endothelial cell membranes and lifting the endothelial cell to enter the sub-endothelial space (Fig.9: Patient 1, Glom_BC_012 (Supp. Movie 5); Patient 2, Glom_BC_053 (Supp. Movie 6); Patient 3, PTCs_BC_001 (Supp. Movie 8)). The contacts were not associated with features of endothelial cell injury. These observations in patients without antibodies against the donor organ set the scene for comparison with patients with antibodies.

### Endothelial cells as antigen-presenting cells

Kidney endothelial cells are considered semi-professional antigen-presenting cells (APC) capable of activating antigen-experienced T cells in the context of alloimmunity (Amersfoort et al., 2022). ‘Missing self’ on the donor’s endothelial cell surface may induce recipient NK cell activation in the kidney microvasculature (Koenig et al., 2019). The outcome of immunological synapse formation may be activatory or immunomodulatory, through exchange of proteins from one cell to the other, a potential mechanism of tolerance induction. The presence of immune synapses between APCs and T or NK cells can be inferred computationally using confocal imaging of human tissue, but only in a small proportion of immune cell pairs where multicellular aggregates are not interfering with the imaging (Wood-Trageser et al., 2023). The extent to which endothelial cells act as APC in vivo, in a human allograft in the context of health or disease, has not been documented. We observed in one instance (in patient 1) a mononuclear cell and an adjacent endothelial cell forming a ‘synapse’: a short stretch of close apposition delineating a space within which fine cellular projections were noted (Fig.9: Patient 1, Glom_BC_001 (Supp. Movie 4)). This was not associated with features of cell injury within either cell.

### Platelets in patient transplant biopsies

Platelets (PLT) play a primary role in haemostasis, aggregating on contact with injured endothelium. More recently, interactions between PLT and immune cells leading to modulation of immune responses have been described using in vitro and animal models, including in glomerular inflammation, where platelets induce leukocyte recruitment (Hoeft et al., 2023; Kuligowski et al., 2006; Scherlinger et al., 2023). Interactions between PLT, and between PLT and the endothelium and/ or immune cells, cannot be resolved in human tissues using light microscopy, single cell RNAseq or spatial transcriptomics. Likewise, single 2D EM sections through PLT cannot reliably document interactions that occur over a small volume in 3D or provide sufficient context for quantification. The nanopathology pipeline thus presents a unique opportunity to study PLT in transplantation and in human glomerular disease. In all three patients, PLT were noted within glomerular capillary loops, in variable abundance, not obviously related to circulating platelet counts. PLT established numerous contacts with each other, forming intracapillary ‘chains’ (Fig.9: Patient 3, Glom_BC_002 (Supp. Movie 9)). A majority of the PLT observed also extended fine cellular projections to establish contact with the endothelial cell surface, and occasionally also with other blood cells. In patient 2, PLT were particularly numerous and, in this patient, an intracapillary megakaryocyte (MGK) was present that appeared to be budding PLT (Fig.9: Patient 2, Glom_BC_103 (Supp. Movie 7)). Peripheral organ (i.e. not bone marrow) MGK may participate in immunosurveillance (lung (Lefrançais et al., 2017)) or produce platelets with different immunological attributes, such as the ability to induce netosis (spleen, (Scherlinger et al., 2023; Valet et al., 2022)) or T cell activation (Pariser et al., 2021).

## Discussion

Human pathologies have clinical features that are visible at different scales: Medical imaging detects features of disease initiation and progression at low resolution in the living intact human body; histopathology detects features at intermediate resolution in tissue removed from the body; and nanopathology detects features in further dissected tissue at high resolution. As resolution increases, so the volume of tissue imaged drops dramatically, and context from the surrounding tissues is lost.

Our newly developed nanopathology pipeline takes the first steps in addressing this challenge, by preserving patient tissue in a state suitable for imaging across scales, and linking a series of imaging modalities that operate at different scales with key sample preparation and analysis steps. We applied the nanopathology pipeline to detect previously unreported features of pathology in a critical clinical challenge - that of immune-mediated injury in patients who have received a kidney transplant.

Human kidney biopsies were fixed in clinic using a protocol that enabled downstream molecular and ultrastructural imaging. Whole biopsies were imaged in minutes, fixed and hydrated, using HiTT, a new synchrotron-based X-ray imaging technique. Immune cells were labelled by non-permeabilisation immunofluorescence and imaged using light microscopy, allowing unequivocal identification of immune cell type in human tissue without disrupting ultrastructure. Features of interest were targeted using the X-ray and light microscopy data, and imaged using volume EM, revealing nanoscale cellular interactions in three dimensions. To image the nanoscale features of an entire kidney biopsy core with volume EM alone would take around 50 years with today’s technology. This nanopathology pipeline reduces that time to around 50 days, focusing on high resolution features of interest in small ROI volumes whilst maintaining mid-resolution structure through the whole biopsy for context.

The nanopathology pipeline was applied to three human transplant biopsies without antibody-mediated rejection, revealing evidence of extensive patrolling of the endothelial cell surface by neutrophils and cells of the monocyte/ macrophage lineage. An immunological synapse between an intraluminal immune cell and an endothelial cell, and platelet interactions with each other and with endothelial cells were noted. Whereas patrolling monocytes have been observed in animal models using intravital imaging, to our knowledge this is the first observation of three-dimensional immune cell behaviours in human tissue from patients.

Our initial analysis of three transplant biopsies has already led to qualitative insights into interactions between circulating cells and the graft endothelium, not previously observed in the human kidney. To derive quantitative data from a larger number of samples, higher throughput is required. In the next iteration of the pipeline, a target of 5 days for multimodal imaging of a biopsy would reduce the time taken by another order of magnitude, significantly increasing the speed of clinical research and bringing multiscale multimodal imaging of human tissue within reach of diagnostic timescales. A number of technical and workflow advances would be needed to reach this target.

HiTT imaging is so fast that it is not a bottleneck in the pipeline, but shipping to and from the synchrotron took around a week for each return journey. Establishment of HiTT at geographically distributed synchrotrons would help to reduce shipping times, especially if co-located with clinical research teams in the same country. In parallel, research groups are working to develop laboratory-based phase contrast X-ray microscopes that would be capable of generating images similar to HiTT from unstained soft tissues, though not as fast (Esposito et al., 2025; William et al., 2022).

The speed of light microscopy is not limiting in the pipeline, but could be improved by decreasing the resolution with which the vibratome slices are imaged dependent on the size of the features of interest, or possibly by using emerging techniques such as light field microscopy (Kim, 2022) or lattice light sheet microscopy (Chen et al., 2014).

The speed of volume EM could be increased by further development of smart workflows that image at high resolution in ROIs and at low resolution everywhere else to maintain context. In this study, we selected ROIs of around 1 x 10^6^ µm^3^, encompassing glomeruli and matching volumes of PTCs. In the next iteration, finer targeting of smaller ROIs with flexible shapes drawn closer to ROI boundaries would increase speed, as would targeting of individual immune cells for high resolution imaging. Finer targeting will require detection of smaller structures and individual cells in the X-ray and light microscopy data, and precise translation of targeting coordinates across imaging modalities. Experimental determination of the optimal resolution for imaging within and outside of ROIs will also be key and will likely depend on the size and composition of the three-dimensional nanoscale pathological features that are identified by the pipeline. High precision smart imaging workflows could also be combined with more sensitive electron detectors and/ or massively parallel imaging systems as a brute force method for faster electron imaging (Eberle et al., 2015; Kievits et al., 2024).

The efficiency, speed, and reproducibility of the overall data analysis workflow could be increased by utilising distributed computing methods, for example Dask (Dask Development Team, 2016) or Nextflow (Di Tommaso et al., 2017) for efficient orchestration of parallel processing workloads across many CPUs and GPUs, in high performance compute or cloud environments. The use of next generation file formats (NGFF; (Moore et al., 2021)) in our pipeline is an important step for integration of future developments that holds the potential to also speed up individual processing operations.

Data analysis could be made more efficient and automated by integrating machine learning algorithms. In the current pipeline, detection, classification and segmentation of glomeruli and peritubular capillaries was performed manually. Advances in deep learning models for computer vision offer several opportunities to automate these processes. Prompt-based foundation models for image segmentation (Kirillov et al., 2023; Ravi et al., 2024) have recently demonstrated promising results when applied to microscopy images including EM (Archit et al., 2025) and have the potential to massively alleviate the manual effort required to segment immune cells. Supervised deep learning-based object detectors (Zou et al., 2023) are also likely worth investigating considering their success in automating analysis in a wide range of EM-related applications, from transmission EM (Su et al., 2023) to cryo-electron tomography (Genthe et al., 2023) and cryo-electron crystallography (Yonekura et al., 2021). Although they could potentially open the path to fully automated identification of immune cells, training object detectors requires the availability of high-quality annotated example data, which is in itself a challenge. Automating analysis of datasets from the nanopathology pipeline would not only increase speed and throughput in the long run, it would also deliver new biological and clinical insights since the data is too rich to be fully mined by human effort alone.

We expect that the nanopathology pipeline will be applicable to other pathologies, within and beyond kidney transplants, likely with adaptations depending on the tissue type, and the ROI size and structure. The main adaptations are likely to be the slicing strategy and the choice and availability of suitable immunolabels able to access the human antigens of interest without permeabilisation. It will also be of great interest to test the nanopathology pipeline in a pathology for which mid-to-high resolution medical imaging has been performed in vivo (e.g. MRI or CT), to layer whole human image data into the pipeline for pathologies where this is commonly used. Doing so may have the potential to link new nanoscale features of disease progression to gross anatomical changes that can be detected in vivo, with the aim of bypassing invasive biopsy sampling procedures.

At the outset of this study, we were unable to find any accessible biobanks that held tissue compatible with the nanopathology pipeline. Having found that fixation in 4% FA and storage in 1% FA at 4°C can preserve the molecular and structural integrity of human tissue for prolonged periods (at least a year based on samples processed through the pipeline to date), we propose that a new form of biobanking might be feasible. Nanopathology biobanks would allow clinical research to leverage all forms of high resolution volume imaging, incorporating not only HiTT and volume EM, but also precise protein localisation using super resolution light microscopy, spatial gene expression at the nanoscale, spatial elemental analysis using nanoSIMS (Jiang et al., 2016) and possibly *in situ* structural biology (McCafferty et al., 2024), if crosslinking macromolecular structures by FA fixation does not render the structures unrecognisable at the Angstrom scale. Samples in nanopathology biobanks could be accrued over a period of time, and optimal sample sets triaged/ selected for downstream analysis by clinical researchers, adding value to individual studies as well as large consortium efforts such as the Human Cell Atlas (https://www.humancellatlas.org/) and the Human Organ Atlas (https://human-organ-atlas.esrf.fr/).

Finally, although developed for research applications we note that, as well as targeted molecular and ultrastructural imaging in regions of interest at nanometer resolution, the nanopathology pipeline captures image data from 100% of the tissue sample at micron resolution. This ‘smart’ approach to tissue imaging and molecular characterisation is in stark contrast to the current standard diagnostic techniques, where most of the diagnostic sample is either left behind in the paraffin block, or trimmed off and lost during sample preparation, with only a small amount of the sample (estimated ∼2% in the case of a kidney biopsy) used for diagnosis. In addition, the observation that immune cells identified using high resolution ultrastructural imaging can also be detected using faster, lower resolution microstructural imaging techniques raises the possibility of integration into future diagnostic practice. The nanopathology pipeline therefore has potential to contribute to new diagnostic histopathology protocols, if the strict turn-around-time requirements in a diagnostic lab can be achieved.

## Materials and Methods

### Sample collection, fixation, division and storage

Material excess to diagnostic needs from indication kidney transplant biopsy samples were taken at Imperial College NHS Trust Renal Unit. Imperial College Healthcare NHS Trust Tissue Bank has ethical approval to both collect human tissue excess to diagnostic needs and release material to researchers (MREC 22/WA/2836). Patients undergoing a procedure at Imperial College Healthcare NHS Trust were asked at time of the procedure if they consented to material surplus to diagnostic needs from their tissues being used for research, using patient information sheet (PIS) v.8. Patient responses were recorded in the electronic patient record and on the pathology request form. Approval for our specific project was registered with Imperial College Healthcare Tissue Bank as project R14094: Outcome analysis after renal transplantation in patients with de novo donor-specific antibodies.

A small piece of the biopsy tissue from each patient was fixed in 4% EM-grade formaldehyde (FA) (TAAB) in 0.1 M phosphate buffer (PB) pH 7.4. Samples were transferred in fixative to the local diagnostic EM facility and left overnight at room temperature to ensure inactivation of any unknown pathogens. The samples were then stored at 4°C until diagnosis was complete. If not needed for diagnosis, the samples were pseudonymised and transported to the Francis Crick Institute.

The database linking NHS sample number and research sample pseudonym was stored in a single secure location within the NHS IT system. Following transportation to the Francis Crick Institute, biopsy samples were logged into a Laboratory Information Management System. Details including the biopsy pseudonym, storage location, and physical status were recorded for each biopsy. Samples were transferred to 1% FA in 0.1 M PB and this storage fixative was replaced approximately every 3 months.

The detailed protocol for biopsy fixation compatible with the nanopathology pipeline is available at protocols.io (dx.doi.org/10.17504/protocols.io.dm6gpzxn8lzp/v1).

### High Throughput Tomography (HiTT) of fixed hydrated biopsies

Fixed hydrated biopsies were washed 3 times in 0.1 M PB and imaged using a stereo microscope (M205 C; Leica Microsystems). Biopsies longer than 6 mm were cut into two pieces using a fresh number 11 surgical scalpel blade and then returned to 1% FA in 0.1 M PB for storage. Cuts through regions that appeared to be cortex were avoided where possible.

Fixed hydrated biopsies were transported by courier to EMBL Hamburg for X-ray imaging using High Throughput Tomography (HiTT) (Albers et al., 2024) on the P14 beamline at the PETRA III synchrotron on the DESY campus. Biopsies were transported in sterile 0.5 ml microcentrifuge tubes (Eppendorf™) filled with 1% FA in 0.1 M PB and sealed with Parafilm^TM^ in temperature-controlled shipping containers (2 to 8°C).

At the beamline, the biopsies were transferred to 10 μl sterile pipette tips (Starlab, Germany) (the tip of which had been sealed using heat) filled with 1% FA in 0.1M PB. The tips containing the tissue were centrifuged at 700 x g for 5 s to remove air bubbles and help the tissue settle towards the bottom of the pipette tip. The open top of the pipette tip was then attached to a metal spine pin with a magnetic base (Mitegen, Ithaca, USA). Sample translation and rotation were controlled using a modified MD3 diffractometer (Arinax, Moirans, France (Cipriani et al., 2007)) with a magnetic base that held the sample in place. This also facilitated the use of the automated sample changer (MARVIN), which allowed pre-mounting of 170 samples at a time and allowed sample changes without re-entering the experimental hutch.

HiTT images of the fixed hydrated biopsies were captured at 18 keV using the 10x objective, resulting in a pixel size of 0.65 µm with a 1250 x 1250 µm field of view (FOV). For phase contrast tomographic imaging, four single tomograms at increasing sample-to-detector distances of 153, 157, 163 and 172 mm were acquired. For each distance, 1810 projections over a 181° rotation angle were recorded, resulting in a total exposure time of 72.4 s for the four distances of each individual scan. Biopsies larger than 1.25 mm long did not fit into the FOV of a single HiTT scan. To resolve this, tiled scans were acquired along the length of each biopsy (usually 5 to 7 scans depending on the length of the tissue) with an overlap of 450 µm, which were then stitched together post-acquisition.

After HiTT imaging, biopsies were transferred back into sterile 0.5 ml microcentrifuge tubes (Eppendorf™) filled with 1% FA in 0.1 M PB and sealed with Parafilm^TM^ for temperature-controlled shipping (2 to 8°C).

### HiTT data processing

After successful completion of data collection, the 3D reconstruction pipeline was automatically triggered. The pipeline consisted of an application written in-house, TOMO CTF (C++), which performs phase-retrieval according to a slightly modified contrast transfer function (CTF) algorithm (Cloetens et al.) as well as tomographic reconstruction using the GridRec algorithm. TOMO CTF was run on the EMBL Hamburg beamline HPC cluster utilising 608 CPUs simultaneously. This resulted in a CTF computing time and reconstruction of 30 secs for each dataset, which could then be viewed instantly using the in-house developed TOMO VIEWER. Each individually reconstructed dataset was output as a series of 2048 tiff images in a 32-bit float format with a nominal pixel size of 0.65 µm.

For biopsies that were imaged with tiled scans, the series of tiled scans were reconstructed and then stitched together into one dataset using NRStitcher (Miettinen et al., 2019).

NRStitcher performed a rigid transformation based on the acquisition coordinates and output a single dataset in.raw file format encompassing the whole kidney biopsy. Image stacks were converted into movies using a custom python script with ffmpeg codecs to facilitate rapid exploration of the microanatomy of the biopsies.

### HiTT data analysis

Total biopsy volumes and surface areas were calculated using a label analysis in Amira (v2021.1, Thermo Scientific). Glomeruli were identified, classified and manually segmented in Amira using a graphics tablet. Volume renderings of glomeruli were visualised in VGStudio MAX (v2024.4, Volume Graphics).

### Vibratome sectioning

Kidney biopsies were embedded vertically in 3% low melting point agarose (Sigma Aldrich) in 0.1 M PB and 60 µm transverse vibratome slices were cut using a VT1200 vibrating blade microtome (Leica Biosystems). Vibratome settings were set to 0.3 mm/s speed of advance and 0.7 mm amplitude, and the blade was made of carbon steel (FEATHER®). Slices were collected and transferred in sequence to an optical bottom 24-well µ-Plate (Ibidi) filled with 0.1 M PB. Vibratome slices were stored in 1% FA in 0.1 M PB at 4°C.

### Autofluorescence imaging

Widefield autofluorescence imaging of vibratome slices was performed using an AxioObserver 7 LSM900 light microscope with Zen software (Version 3.10; Carl Zeiss Ltd). The autofluorescence signal was imaged using the preset EGFP-channel illumination and detection settings. To locate slices, a 2D tile scan covering the entire 24-well plate was acquired using a 2.5x/ 0.085 NA objective. Stage speed and acceleration were reduced to 1% to prevent sample drift. Higher magnification imaging of each slice was then performed using a 20x/ 0.8 NA objective. Images were acquired as a coarse z-stack (10 µm interval) and 2×2 tile scan. The structures observed in the autofluorescence data (tubules and glomeruli) were then registered with the fixed-hydrated whole biopsy HiTT data to confirm which vibratome slices contained the pre-selected ROIs. One slice from each biopsy (containing a central portion of the selected glomerulus) was then selected to progress through the nanopathology pipeline for molecular and ultrastructural imaging.

### Non-permeabilisation immunofluorescence labelling

All CD16 immunofluorescence labelling and Hoechst staining steps were performed in sterile 24-well plates, at room temperature on a plate shaker, unless otherwise specified. Slices were washed 3 times in 0.1 M PB, then incubated in 0.05 M glycine (Sigma) in 0.1 M PB for 30 min. Slices were washed 3 times in 0.1 M PB, then incubated in a blocking solution of 1% bovine serum albumin (BSA; Sigma Aldrich) plus 0.5 % normal goat serum (Sigma-Aldrich) in 0.1 M PB for 30 min. Slices were incubated in the primary antibody (rabbit monoclonal [EPR22409-124] to CD16 [abcam]) diluted 1:50 in 0.5% BSA in 0.1 M PB overnight at 4°C without agitation. Slices were washed 3 times in 0.1 M PB then incubated in the secondary antibody (donkey anti-rabbit IgG H&L [Alexa Fluor® 647] preadsorbed [abcam]) diluted 1:100 in 0.5% BSA plus 4 µg/ml Hoechst 33342 (Invitrogen Molecular Probes) in 0.1M PB for 90 min. Slices were washed a further 3 times in 0.1 M PB for a total of 30 min. Slices were returned to 1% FA in 0.1 M PB for storage. For patient 3, the first confocal fluorescence dataset was acquired immediately after IF labelling, prior to the slice being returned to 1% FA.

### Confocal fluorescence microscopy

Confocal fluorescence microscopy was performed using an AxioObserver 7 LSM900 light microscope with Zen software (Version 3.10; Carl Zeiss Ltd) and a 40x/ 1.2 NA water immersion lens. Vibratome slices were imaged in 0.1 M PB in a glass bottom petri-dish (MatTek P35G-1.5-14-C). In some cases, vibratome slices were mounted underneath a 13 mm diameter coverslip, using part of a Secure-Seal™ 9 mm diameter spacer (Invitrogen) attached to the glass of the petri-dish to prevent crushing of the tissue. For slices which were not mounted under a coverslip, stage speed and acceleration were reduced to 1% to prevent sample drift. To locate the edges of the slice, a fast widefield montage was acquired using the Hoechst H33342 illumination and detection settings. Smart setup was then used to configure multi-channel confocal fluorescence microscopy settings for AF647 and Hoechst H33342. A tiled region was created to cover the entire slice and up to 50 focus plane support points were added. For each support point position, the lowest plane where signal from the tissue could be seen was measured, and an offset of ∼20 µm was applied. The confocal fluorescence montage was acquired as an ∼55 µm z-stack with an ∼0.5 µm interval and ∼200 nm xy pixel size, using the tile-setup support points to define the centre of each stack. For full volume imaging of the slices (performed for patient 1 and patient 2 biopsies but not patient 3) the vibratome slice was subsequently flipped, re-mounted, and confocal fluorescence microscopy was repeated for the second side.

### Confocal fluorescence data processing

Confocal fluorescence data was stitched using the BigStitcher plugin (Hörl et al., 2019) for the Fiji image processing platform (Schindelin et al., 2012). Data was converted to N5 format (https://github.com/saalfeldlab/n5) during import. Pairwise shifts were calculated using the phase-correlation algorithm, with default settings. The groups of aligned tiles were then assessed and a view where several tiles were well aligned was noted down. Global optimisation was applied using the default settings and ‘fixing’ the group/view previously noted as having several good links. The fully aligned data was exported in 16-bit format using the ‘Image Fusion’ option with linear interpolation and subsequently saved as.tiff stacks.

### Sample processing for volume EM

Vibratome slices were prepared for SBF-SEM using a modified version of the protocol from the National Center for Microscopy and Imaging Research (NCMIR) (Deerinck et al., 2010). Slices were post-fixed in 2.5% glutaraldehyde (GA; EM-grade; TAAB) and 1% FA (EM-grade; TAAB) in 0.1 M PB for 1 h at RT. Slices were washed in 0.1 M PB, stained in 2% osmium tetroxide (TAAB)/ 1.5% potassium ferricyanide (Sigma-Aldrich) for 1 h at 4°C in the dark, washed in purified H_2_O and stored in H_2_O at 4°C overnight. Slices were stained in 1% thiocarbohydrazide (Sigma-Aldrich) for 20 min at RT, washed in H_2_O and then stained in 2% osmium tetroxide (TAAB) for 30 min at RT. Slices were washed in H_2_O, stained in 1% aqueous uranyl acetate (Agar Scientific) overnight at 4°C in the dark, washed again in H_2_O, then incubated for 30 min at 60°C in Walton’s lead aspartate (Walton, 1979) prepared using aspartic acid and lead nitrate (Sigma-Aldrich). Slices were washed in H_2_O then dehydrated through a graded ethanol series for 10 min per step (30%, 50%, 70%, 90%, 100%, 100%). Slices were incubated in 100% propylene oxide (PO; TAAB) for 10 min at RT. Slices were then infiltrated with a 3:1 mixture of PO: Durcupan resin (Sigma-Aldrich) for 2 h at RT, followed by a 1:1 mixture of PO: Durcupan resin for 2 h at RT, and then a 1:3 mixture of PO: Durcupan resin overnight at RT. Slices were further infiltrated with 3 x 2 h changes of 100% Durcupan resin at RT, before ‘flat embedding’ in a thin layer of resin sandwiched between 2 sheets of thermoplastic film (Aclar®). The resin was polymerized at 60°C for 48 h. After resin polymerization, the Aclar® sheets were peeled from the resin and the vibratome slices were cut out using a razor blade.

### X-ray microscopy of resin-embedded vibratome slices

The resin embedded vibratome slices were imaged by X-ray microscopy using either synchrotron-based HiTT (patients 1 & 2) or laboratory-based micro computed tomography (microCT; patient 3).

For HiTT, the sample was first mounted flat on the aluminium pin and trimmed as above. For HiTT imaging, the set up was identical to that described earlier for imaging fixed hydrated biopsies, except for the object-to-detector distances (73, 77, 83 and 92 mm) and the energy (set to 23.5 keV to deliver better contrast from the heavy metal-stained tissue). No tiling was required as the sample was smaller than the field of view, so each vibratome section could be imaged and the data reconstructed in less than 3 minutes.

For microCT, a Versa 510 XRM (Zeiss) was used to image the sample through the thinnest axis of the vibratome slice. A 20x lens was used with a source voltage of 40 kV at 3W with no beam filter. Three-thousand and one projections were acquired over 360° with an exposure time of 15 s per projection. Binning was set at 2 with a voxel size of 0.929 µm with a 925.51 µm² field of view. Projection data was reconstructed using Scout-and-Scan Control System Reconstructor (Zeiss). The sample was then mounted flat on an aluminium pin (for the Gatan 3View system) using CircuitWorks® Conductive Epoxy (Chemtronics). The edges of the samples were trimmed to form a smooth-sided cuboid using glass knives, a diamond knife (trim 90; Diatome) and an EM UC7 ultramicrotome (Leica Microsystems).

### Segmentation of ROIs to drive targeted SBF-SEM

ROIs to be imaged by SBF-SEM were segmented in the XRM data acquired from the resin-embedded vibratome slices. For each vibratome slice, the entire volume containing the glomerulus was segmented alongside a similar volume of an adjacent tubular region. The ROIs were segmented using IMOD software (Mastronarde, 1997) and binary mask image stacks of these segmented volumes were written using the IMOD Model Painter program (https://bio3d.colorado.edu/imod/doc/man/imodmop.html).

### Serial Block Face SEM

SBF-SEM acquisition was performed with a Gatan 3View system mounted on a Zeiss Gemini450 scanning electron microscope. The acquisition was performed using the open source software SBEMimage (Titze et al., 2018).

The pin with the mounted and trimmed sample was loaded into the 3View sample holder and inserted into the microscope stage. Sample approach to the knife was performed at atmospheric pressure with the microscope door open using a cutting thickness of 200 nm and a speed of 0.5 mm/s. When the first sections were cut, the door was closed, and the chamber was pumped to vacuum. The sample was left for 24 h in vacuum before proceeding with the next step to allow for potential movement due to degassing of the sample and stage drift. Typically, this resulted in a few microns of downwards drift, so a second approach of the knife to the sample was performed in vacuum using the ‘approach’ command in SBEMimage with steps of 0.5-1 µm (100 nm slice thickness), whilst monitoring whether the block surface had been cut by acquiring overview images.

As soon as the tissue was exposed at the block surface, the tissue morphology was used to visually register the tissue orientation to the XRM data from the resin embedded slice, allowing the ROI segmentations to be used to create two tile sets in SBEMimage to drive high resolution imaging in those regions.

For imaging, the SEM was operated at 1.5 kV acceleration voltage with a nominal current of 350 pA (patients 1 & 2) or 300 pA (patient 3). Backscattered electrons were collected with a Gatan OnPoint detector (contrast: 99, brightness 15 to 19). To reduce charging artefacts, we used a focal charge compensation device (Zeiss Microsystems) (Deerinck et al., 2017), obtaining a pressure ranging between 1.8×10^-3^ to 2.7×10^-3^ in the microscope chamber. For the ROIs, the acquisition pixel size was 8.24 nm (patient 1) or 10 nm (patient 2 and patient 3) and the tile size 4096 x 3072 pixels, with a dwell time of 1.6 µs. A typical ROI was covered by a set of 5 x 6 tiles, with an overlap of 200 pixels, but the tile activation/ deactivation was optimized during the acquisition to image the entire ROI. After imaging each block surface, 50 nm sections were removed with a cutting speed of 0.2 mm/s and a knife retraction speed of 0.5 mm/s before another imaging iteration took place. Prior to high resolution imaging of the ROIs, a low resolution overview image (pixel size 200-300 nm, dwell time 0.8 µs) was acquired and used for automated debris detection. If a big change in image contrast was detected (a sign of the potential presence of debris on the block surface), the knife was swept once above the block surface, without cutting, to remove the debris, before high resolution imaging started. Focus was optimised for each set of ROI tiles using the SBEMimage function ‘focus tracking’ with manual refocusing.

With this setup, the acquisition of 2 large ROIs (roughly 200 µm x 200 µm on average) spanning the entire thickness of a 50 µm section lasted 2 to 3 weeks and generated 530-620 GB of raw data.

### SBF-SEM data processing

SBF-SEM tiles were aligned and stitched using the TrakEM2 plugin (Cardona et al., 2012) in Fiji (Schindelin et al., 2012). Image data was imported using an ‘imagelist’ text file exported from SBEMimage, which ensured roughly correct initial placement of the tiles in 3D space. The xy pixel size was set according to the acquisition pixel size of the given dataset and the z-scaling was set to the section thickness (50 nm). The brightness and contrast of the data was adjusted using the option ‘set min and max layer-wise’. The values for min and max adjustment were determined by opening a representative image from the raw data in Fiji and finding min and max pixel intensity display values where the histogram filled the range without cutting off extreme values.

Alignment and stitching were performed using ‘Align Multi-Layer Mosaic’ in batches of ∼100 sections to prevent the computation stalling, with ‘tiles are roughly in place’ selected. Settings for intra-layer registration parameters were as follows:

● SIFT Scale Invariant Interest Point Detector (initial gaussian blur = 1.60 px, steps per scale octave = 3, minimum image size = 800 px, maximum image size = 1200 px).
● SIFT Feature Descriptor (feature descriptor size = 4, feature descriptor orientation bins = 8, closest/next closest ratio = 0.92).
● Geometric Consensus Filter (maximal alignment error = 20 px, minimal inlier ratio = 0.0, minimal number of inliers = 5, expected transformation = Translation, ignore constant background = no).
● Alignment parameters (desired transformation = affine, correspondence weight = 1.0, regularize = yes, optimization - maximal iterations = 2000, optimization maximal plateau width = 200, filter outliers = no).
● Regularization parameters (regularizer = translation, lambda = 0.01).

Settings for cross-layer registration parameters were as follows:

● SIFT Scale Invariant Interest Point Detector (initial gaussian blur = 1.60 px, steps per scale octave = 3, minimum image size = 100 px, maximum image size = 500 px).
● SIFT Feature Descriptor (feature descriptor size = 8, feature descriptor orientation bins = 8, closest/next closest ratio = 0.92).
● Geometric Consensus Filter (maximal alignment error = 20 px, minimal inlier ratio = 0.05, minimal number of inliers = 5, expected transformation = Translation, ignore constant background = no).
● Alignment parameters (desired transformation = affine, correspondence weight = 1.0, regularize = yes).
● Regularization parameters (regularizer = translation, lambda = 0.01).

Once the batch of sections was successfully aligned and the new positions for tiles saved, all tiles on slice numbers n-2 and n-1 (with n being the final slice number in the batch) were locked in position. The next batch of slices was then processed using the same method, starting with the 2 ‘locked’ slices, so that there was an overlap of 3 slices between each batch processed. Once all slices were aligned and stitched, the EM data was exported and saved as a tiff stack.

### SBF-SEM data analysis

To determine the volumes of glomeruli and PTCs in the SBF-SEM datasets, the processed (stitched/ aligned) dataset was binned in xy by a factor of 12. Binned data was converted to. mrc file format using IMOD open-source software (Kremer et al., 1996) and the file header metadata was edited to give the correct pixel values in xyz. The data was then segmented using IMOD.

For segmentation of the glomerular tuft, manual annotation with closed contours using the sculpt drawing tool (https://www.andrewnoske.com/student/imod.php) was performed once every 20 slices. Slices that were not manually annotated were segmented using linear interpolation. Once segmentation was complete, a triangular surface mesh was generated from the contours and the volume within the mesh was recorded in µm³.

For segmentation of PTC volumes, a mixture of semi-automated and manual annotation was used. Once every 20 slices, the blood vessel lumen was segmented using the autocontour threshold-based segmentation tool. Manual corrections using the sculpt or join drawing tools were used where necessary. To correct for interpolation errors at points of blood vessel branching, additional key contours were added manually, and interpolated contours were regenerated. Once segmentation was complete, a triangular surface mesh was generated from the contours and the volume within the mesh was recorded in µm³.

For patient 3, confocal fluorescence data was only collected for part of the volume imaged by SBF-SEM. To calculate the volume of glomerular tuft and PTCs imaged using both light and electron microscopy, the glomerular and PTC volumes were first segmented using the binned EM data. The confocal fluorescence data (Hoechst channel) was then transformed to match the binned EM data using the BigWarp plugin in Fiji (Bogovic et al., 2016). The transformed LM data was exported with the same field of view and resolution to match that of the binned EM data. The transformed LM data was then converted to.mrc file format and loaded in IMOD with the glomerular tuft or PTC model created from segmentation of the EM data. Contours corresponding to the section of the model where the LM data was absent were removed and the model was re-saved. A triangular surface mesh was generated from the new model and the volume within the mesh was recorded in µm³.

### Viewing multimodal image data in MoBIE

The Fiji plugin MoBIE (Pape et al., 2023) was used for multimodal image data registration and viewing. For import into MoBIE, datasets were converted to the OME-Zarr file format (Moore et al., 2023). Initial data transformations (to enable multimodal data registration) were performed using the BigWarp plugin (Bogovic et al., 2016).

For registration of autofluorescence and confocal fluorescence data to fixed hydrated biopsy HiTT data, distinctive shapes of the tubules were used as landmarks and light microscopy datasets were transformed using BigWarp’s affine transformation method.

For registration of confocal fluorescence microscopy datasets to XRM (HiTT and microCT) of resin-embedded samples, nuclei were used as landmarks. Where possible, confocal fluorescence data was transformed using BigWarp’s affine algorithm; in some cases, where the glomerulus had shifted independently to the rest of the vibratome slice, the confocal fluorescence data was transformed in Fiji using BigWarp’s thin plate spline algorithm.

For registration of electron microscopy datasets to XRM (microCT and HiTT) of resin-embedded samples, EM data was transformed relying on BigWarp’s similarity transformation option. In cases where imaging the same sample with different imaging modalities resulted in the image data being mirrored, thus causing rigid and similarity alignment transforms to fail, the data was flipped in the z-direction prior to application of the similarity transformation.

### Location of blood cells in multimodal datasets

To locate and annotate the positions of the relevant blood cells (immune cells and platelets/ megakaryocytes) within the SBF-SEM data, the stitched and aligned EM data in OME-Zarr format was loaded into napari (Ahlers et al., 2023). The data was systematically inspected in quadrants to identify all blood cells, and the scattered points tool (size and opacity = max, out of slice = yes) was used to visibly mark the locations of these cells. The points data was saved as a.csv file, ‘to_mask’ was used to create a binary mask of the points, and ‘.scale’ was used to ensure that the pixel scaling of the points mask file matched the pixel scaling of the image data file (https://napari.org/dev/api/napari.layers.Points.html#napari.layers.Points). The points mask data was then exported as a tiff file using ‘tifffile’ (https://github.com/cgohlke/tifffile). The binary mask.tiff was binned in xy by a factor of 8 before conversion to OME-Zarr and import into the MoBIE project. The blood cell locations mask was then transformed using the same matrix used to previously transform the SBF-SEM data so that it aligned with the resin HiTT data. The transformed versions of the SBF-SEM data, confocal fluorescence data, and blood cell locations data (all aligned with each other in the spatial co-ordinates of the un-transformed resin HiTT data) were opened in the MoBIE viewer. The blood cell locations mask provided a visible marker and enabled efficient re-identification of the blood cells within the registered multimodal image datasets. For each cell, a zoomed-in view of its location was saved within the MoBIE interface and named with a unique cell identifier number.

## Data Availability

All data produced in the present study are available upon reasonable request to the authors and will be made available in public image archives on journal publication

## Acknowledgements

Human samples used in this research project were obtained from the Imperial College Healthcare Tissue Bank (ICHTB). ICHTB is supported by the National Institute for Health Research (NIHR) Biomedical Research Centre based at Imperial College Healthcare NHS Trust and Imperial College London. ICHTB is approved by Wales REC3 to release human material for research (22/WA/2836). EMBL ethical approval for the Schwab/ Duke/ Uhlmann project: A Nanopathology Platform for the prediction and early detection of disease in kidney transplant rejection (BIAC 2022-032) was granted in December 2022.

We acknowledge funding for staff and resources from the UKRI Medical Research Council (grant number MR/W031426/1), which supported Alana Burrell, Matthew Lawson, and Xiaofan Xiong. This work was supported by the Francis Crick Institute which receives its core funding from Cancer Research UK (CC1076, CC1295), the UK Medical Research Council (CC1076, CC1295), and the Wellcome Trust (CC1076, CC1295). Candice Roufosse is supported by the NIHR Imperial Biomedical Research Centre (BRC). Matthew Hartley is supported by internal funding from the European Molecular Biology Laboratory. Virginie Uhlmann is supported by internal funding from the European Molecular Biology Laboratory and the University of Zurich. Infrastructure support for this research was provided by the NIHR Imperial Biomedical Research Centre (BRC).

We thank the Francis Crick Institute Electron Microscopy Science Technology Platform team for support, and the Electron Microscopy Unit at North West London Pathology under the leadership of Linda Moran for assistance with sample handling. We acknowledge the help and support of Ksenia Denisova, Fabio De Marco and Angelika Svetlove of the Hamburg X-ray Imaging Team along with wider members of the EMBL Hamburg Community for support and assistance in the acquisition and processing of the HiTT X-ray data.

For the purpose of open access, the author has applied a Creative Commons Attribution (CC BY) license to any Author Accepted Manuscript version arising.

