## Supplementary Material for "A nanopathology pipeline for clinical research across scales using human tissue"

#### Supplementary Tables

**Supplementary Table 1. Manual quantification of Intraluminal blood cells (BC) in ROIs.** ROIs are 1) single glomerulus, all tuft intracapillary volume in the 60 micron-thick vibratome slice (Glom); 2) for peritubular capillaries (PTCs), all PTC intraluminal volume in the 60 micron-thick vibratome slice. The table has one line for each glomerular or PTC blood cell counted (in order of cell visualisation, scrolling from top to bottom of slice).

| Patient 1 (EM4652) Polyoma virus nephropathy | Morphology | CD16 status |
|---|---|---|
| **Glomerulus** | | |
| Glom_BC_001 | Mononuclear_Small | pos |
| Glom_BC_002 | Mononuclear_Small | pos |
| Glom_BC_003 | Mononuclear_Large | neg |
| Glom_BC_004 | Mononuclear_Large | neg |
| Glom_BC_005 | PLT | neg |
| Glom_BC_006 | Polymorph? | pos |
| Glom_BC_007 | PLT | neg |
| Glom_BC_008 | PLT | neg |
| Glom_BC_009 | PLT | neg |
| Glom_BC_010 | Mononucelar_Small | neg |
| Glom_BC_011 | Mononuclear_Large | weak pos |
| Glom_BC_012 | Mononucelar_Small | pos |

| | | |
|---|---|---|
| Glom_BC_013 | PLT | neg |
| Glom_BC_014 | PLT | neg |
| Glom_BC_015 | Mononucelar_Small | weak pos |
| Glom_BC_016 | PLT | neg |
| Glom_BC_017 | Mononucelar_Small | pos |
| Peritubular capillaries | | |
| PTCs_BC_001 | Mononucelar_Small | pos |
| PTCs_BC_002 | Mononucelar_Small | pos |
| PTCs_BC_003 | Mononucelar_Small | neg |
| PTCs_BC_004 | PLT | neg |
| PTCs_BC_005 | Mononucelar_Small | neg |
| PTCs_BC_006 | PLT | neg |
| PTCs_BC_007 | PLT | neg |
| PTCs_BC_008 | PLT | neg |
| PTCs_BC_009 | Mononucelar_Small | pos |
| PTCs_BC_010 | Mononucelar_Small | pos |
| PTCs_BC_011 | Mononucelar_Small | neg |
| Patient 2 (EM4654)<br>Acute tubular injury | Morphology | CD16 status |
| Glomerulus | | |
| Glom_BC_001 | PLT | neg |
| Glom_BC_002 | PLT | neg |
| Glom_BC_003 | Mononucelar_Small | neg |
| Glom_BC_004 | Mononuclear_Small | neg |
| Glom_BC_005 | PLT | neg |
| Glom_BC_006 | PLT | neg |
| Glom_BC_007 | PLT | neg |
| Glom_BC_008 | PLT | neg |

| | | |
|---|---|---|
| Glom_BC_009 | PLT | neg |
| Glom_BC_010 | PLT | neg |
| Glom_BC_011 | Polymorph | pos |
| Glom_BC_012 | PLT | neg |
| Glom_BC_013 | PLT | neg |
| Glom_BC_014 | Mononuclear_Large | pos |
| Glom_BC_015 | PLT | neg |
| Glom_BC_016 | PLT | neg |
| Glom_BC_017 | PLT | neg |
| Glom_BC_018 | PLT | neg |
| Glom_BC_019 | PLT | neg |
| Glom_BC_020 | PLT | neg |
| Glom_BC_021 | PLT | neg |
| Glom_BC_022 | PLT | neg |
| Glom_BC_023 | PLT | neg |
| Glom_BC_024 | PLT | neg |
| Glom_BC_025 | PLT | neg |
| Glom_BC_026 | PLT | neg |
| Glom_BC_027 | PLT | neg |
| Glom_BC_028 | PLT | neg |
| Glom_BC_029 | PLT | neg |
| Glom_BC_030 | PLT | neg |
| Glom_BC_031 | Mononuclear_Small | weak pos |
| Glom_BC_032 | Mononuclear_Small | weak pos |
| Glom_BC_033 | PLT | neg |
| Glom_BC_034 | PLT | neg |
| Glom_BC_035 | PLT | neg |
| Glom_BC_036 | PLT | NEG |
| Glom_BC_037 | Mononuclear_Large | weak pos |

| | | |
|---|---|---|
| Glom_BC_038 | PLT | neg |
| Glom_BC_039 | PLT | neg |
| Glom_BC_040 | Polymorph | weak pos |
| Glom_BC_041 | Polymorph | neg |
| Glom_BC_042 | PLT | neg |
| Glom_BC_043 | PLT | neg |
| Glom_BC_044 | PLT | neg |
| Glom_BC_045 | PLT | neg |
| Glom_BC_046 | PLT | neg |
| Glom_BC_047 | Polymorph | weak pos |
| Glom_BC_048 | PLT | neg |
| Glom_BC_049 | PLT | neg |
| Glom_BC_050 | PLT | neg |
| Glom_BC_051 | PLT | neg |
| Glom_BC_052 | PLT | neg |
| Glom_BC_053 | Mononuclear_Small | neg |
| Glom_BC_054 | PLT | neg |
| Glom_BC_055 | PLT | neg |
| Glom_BC_056 | Mononuclear_Small | neg |
| Glom_BC_057 | PLT | neg |
| Glom_BC_058 | PLT | neg |
| Glom_BC_059 | PLT | neg |
| Glom_BC_060 | PLT | neg |
| Glom_BC_061 | PLT | neg |
| Glom_BC_062 | PLT | neg |
| Glom_BC_063 | Polymorph | weak pos |
| Glom_BC_064 | PLT | neg |
| Glom_BC_065 | Mononuclear_Small | weak pos |
| Glom_BC_066 | PLT | neg |

| | | |
|---|---|---|
| Glom_BC_067 | PLT | neg |
| Glom_BC_068 | PLT | neg |
| Glom_BC_069 | PLT | neg |
| Glom_BC_070 | PLT | neg |
| Glom_BC_071 | PLT | neg |
| Glom_BC_072 | PLT | neg |
| Glom_BC_073 | PLT | neg |
| Glom_BC_074 | PLT | neg |
| Glom_BC_075 | PLT | neg |
| Glom_BC_076 | PLT | neg |
| Glom_BC_077 | Mononuclear_Large | weak pos |
| Glom_BC_078 | Polymorph | weak pos |
| Glom_BC_079 | PLT | neg |
| Glom_BC_080 | PLT | neg |
| Glom_BC_081 | Mononuclear_Large | neg |
| Glom_BC_082 | PLT | neg |
| Glom_BC_083 | PLT | neg |
| Glom_BC_084 | PLT | neg |
| Glom_BC_085 | Mononuclear_Small | weak pos |
| Glom_BC_086 | PLT | neg |
| Glom_BC_087 | PLT | neg |
| Glom_BC_088 | Polymorph | neg |
| Glom_BC_089 | Mononuclear_Small | neg |
| Glom_BC_090 | PLT | neg |
| Glom_BC_091 | PLT | neg |
| Glom_BC_092 | PLT | neg |
| Glom_BC_093 | PLT | neg |
| Glom_BC_094 | PLT | neg |
| Glom_BC_095 | PLT | neg |

| | | |
|---|---|---|
| Glom_BC_096 | Mononuclear_Small | pos |
| Glom_BC_097 | PLT | neg |
| Glom_BC_098 | Polymorph | weak pos |
| Glom_BC_099 | PLT | neg |
| Glom_BC_100 | PLT | neg |
| Glom_BC_101 | PLT | neg |
| Glom_BC_102 | PLT | neg |
| Glom_BC_103 | Mononuclear_Very_large | neg |
| Glom_BC_104 | PLT | neg |
| Glom_BC_105 | PLT | neg |
| Glom_BC_106 | Hoechst+ Blob | neg |
| Glom_BC_107 | PLT | neg |
| Glom_BC_108 | PLT | neg |
| Glom_BC_109 | PLT | neg |
| Glom_BC_110 | PLT | neg |
| Glom_BC_111 | Polymorph | pos |
| Glom_BC_112 | Mononuclear_Small | pos |
| Peritubular capillaries | | |
| PTCs_BC_001 | PLT | neg |
| PTCs_BC_002 | PLT | neg |
| PTCs_BC_003 | PLT | neg |
| PTCs_BC_004 | PLT | neg |
| PTCs_BC_005 | PLT | neg |
| PTCs_BC_006 | PLT | neg |
| Patient 3 (EM4629) Borderline for T cell-mediated rejection | Morphology | CD16 status |

| Glomerulus | | |
|---|---|---|
| Glom_BC_001 | Polymorph | pos |
| Glom_BC_002 | PLT | |
| Glom_BC_003 | Mononuclear_Large | pos |
| Glom_BC_004 | PLT | |
| Glom_BC_005 | PLT | |
| Glom_BC_006 | Mononuclear_large | neg |
| Glom_BC_007 | PLT | |
| Glom_BC_008 | PLT | |
| Glom_BC_009 | Mononuclear_Small | pos |
| Glom_BC_010 | PLT | |
| Glom_BC_011 | PLT | |
| Glom_BC_012 | PLT | |
| Glom_BC_013 | Mononuclear_Small | pos |
| Glom_BC_014 | PLT | |
| Glom_BC_015 | PLT | |
| Glom_BC_016 | PLT | |
| Glom_BC_017 | PLT | |
| Glom_BC_018 | Granulocyte no nucleus | neg |
| Glom_BC_019 | Mononuclear_Small | pos |
| Glom_BC_020 | PLT | |
| Glom_BC_021 | PLT | |
| Glom_BC_022 | PLT | |
| Glom_BC_023 | PLT | |
| Glom_BC_024 | Mononuclear_Small | neg |
| Glom_BC_025 | PLT | |
| Glom_BC_026 | Mononuclear_Small | pos |
| Glom_BC_027 | Mononuclear_Large | n/a |
| Glom_BC_028 | Polymorph | n/a |

| | | |
|---|---|---|
| Glom_BC_029 | PLT | n/a |
| Glom_BC_030 | PLT | |
| Peritubular capillaries | | |
| PTCs_BC_001 | Mononuclear_Small | pos |
| PTCs_BC_002 | Mononuclear_Small | neg |

#### Supplementary Figure Legends

**Supplementary Figure 1. Modification of diagnostic pipeline.** In clinical practice, when a transplant biopsy is indicated, 2 to 3 cores of the transplanted kidney are taken under ultrasound guidance. In order to provide material for all techniques needed to reach a diagnosis, the cores are ideally split at bedside to deliver portions of the material for light microscopy, immunofluorescence and electron microscopy (EM). Exact clinical practice varies depending on the centre, the amount of tissue obtained for each patient (some procedures yielding more tissue than others) and the clinical features. In our centre, the sample for EM is placed in glutaraldehyde. In order to preserve both ultrastructure and molecular antigenicity in a format compatible with clinical research as well as diagnostic EM, a modification was introduced whereby, if possible (as determined by the clinician at bedside), a piece of the biopsy between 3 and 10 mm long that would have been fixed using glutaraldehyde was instead fixed in 4% EM-grade formaldehyde (FA) in 0.1 M phosphate buffer (PB). If needed for diagnosis, the sample was transferred to glutaraldehyde and processed as usual. If not needed for diagnosis, the biopsy tissue was released for research and transferred to 1% FA in 0.1M PB for long term storage at 4°C.

**Supplementary Figure 2. Examples of open, segmentally sclerosed and globally sclerosed glomeruli imaged using HiTT.** The top three images show three orthoslices from an 'open' glomerular tuft, with visible blood vessel lumina containing red blood cells. The centre three images show three orthoslices from a segmentally sclerosed glomerulus. Here the glomerular tuft is mostly 'closed' due to sclerotic tissue replacing the majority of the capillaries, with a few functional open capillaries remaining. The bottom three images show three orthoslices from a globally sclerosed glomerulus. There are no visible capillaries indicating that the glomerular unit is no longer functional. The globally sclerosed glomerulus is smaller compared to the open and segmentally sclerosed glomeruli.

**Supplementary Figure 3. Targeting strategy for molecular and ultrastructural imaging (patients 1 & 3).** A-E) patient 1. A) Transverse orthoslice through the reconstructed HiTT tomogram showing a glomerulus (box) selected for downstream molecular and ultrastructural imaging. B) Selected glomerulus (box) displayed in the longitudinal plane of the biopsy and C) zoomed out to show the selected glomerulus in the context of the whole biopsy. D) Widefield autofluorescence image of vibratome slice 15 containing a large part of the selected glomerulus. E) Overlay of the widefield autofluorescence data (green) onto the fixed hydrated biopsy HiTT data (greyscale) for vibratome slice 15. F-J) patient 3. F) Transverse orthoslice through the reconstructed HiTT tomogram showing a glomerulus (box) selected for downstream molecular and ultrastructural imaging. G) Selected glomerulus (box) displayed in the longitudinal plane of the biopsy and H) zoomed out to show the selected glomerulus in the context of the whole biopsy. I) Widefield autofluorescence image of vibratome slice 18 containing a large part of the selected glomerulus. J) Overlay of the widefield autofluorescence data (green) onto the fixed hydrated biopsy HiTT data (greyscale) for vibratome slice 18.

**Supplementary Figure 4. Molecular imaging using non-permeabilisation immunofluorescence (patient 1).** A) Confocal fluorescence image of vibratome slice 15 showing CD16-AF647 (red) and Hoechst H33342 (blue). B) Cluster of CD16-positive immune cells in a peritubular region. C) CD16-positive immune cell in a glomerulus. D) Confocal fluorescence data from slice 15 registered to the fixed hydrated biopsy HiTT data (confocal slice is shown perpendicular to the original imaging plane). The target glomerulus is highlighted (box). The target glomerulus is shown in the fixed hydrated biopsy HiTT data (E), confocal fluorescence data (F), and in the overlay of fluorescence onto HiTT data (G).

**Supplementary Figure 5. Molecular imaging using non-permeabilisation immunofluorescence (patient 3).** A) Confocal fluorescence image of vibratome slice 18 showing CD16-AF647 (red) and Hoechst H33342 (blue). B) CD16-positive immune cells in a peritubular region. C) CD16-positive immune cell in a glomerulus. D) Confocal fluorescence data from slice 18 registered to the fixed hydrated biopsy HiTT data (confocal slice is shown perpendicular to the original imaging plane). The target glomerulus is highlighted (box). E-G) 'En face' images are shown rather than perpendicular (as shown in Fig.4G,H and Supp.Fig.4F,G) because the selected vibratome slice was partially outside of the original fixed hydrated HiTT scan, leading to empty (black) areas in the perpendicular plane of the registered data. The target glomerulus is shown in the fixed hydrated biopsy HiTT data (E), confocal fluorescence data (F), and in the overlay of fluorescence onto HiTT data (G).

**Supplementary Figure 6**. **Post-embedding X-ray microscopy (XRM) of heavy metal stained, resin embedded vibratome slices (patients 1 & 3).** A-C) Patient 1 slice, imaged using HiTT. A) An '*en face*' 2D plane from the 3D tomogram of the vibratome slice showing key structures including a glomerulus and peritubular capillaries. B) A side view of the same 60 µm thick slice. C) The increased contrast in the samples delivered improved contrast and resolution in the HiTT datasets. The glomerulus is outlined with an orange dashed line and example tubules are outlined in purple. Nuclei of the tubular epithelium cells (green arrows) are clearly visible. D-F) Patient 3 slice, imaged using a Versa 510 lab-based microCT**.** D) An '*en face*' 2D plane from the 3D tomogram of the vibratome slice showing key structures including a glomerulus and peritubular capillaries. E) A side view of the same 60 µm thick slice. F) The increased contrast in the samples delivered improved contrast and resolution in the HiTT datasets. The glomerulus is outlined with an orange dashed line and example tubules are outlined in purple. Red blood cells in the glomerular capillary loops (red arrows) and nuclei of the tubular epithelium cells (green arrows) are clearly visible.

**Supplementary Figure 7. Resin-HiTT-targeted SBF-SEM imaging of a glomerulus ROI and a PTC-rich ROI (patient 1).** A) 3D visualization of the HiTT data acquired from the heavy metal stained, resin embedded vibratome slice. Two regions of interest (ROIs) containing a glomerulus and a region rich in peritubular capillaries were identified and segmented (green) in the HiTT data. B) Low resolution overview image of the block surface acquired using SBF-SEM. The two ROIs identified in the HiTT volume are shown (green masks). C) Screenshot of the SBEMimage interface showing a grid that covers the entire imaging field. Active tiles (green) differ for each block surface exposed during cutting according to the size and position of the ROIs. The crosshair (red) shows the current stage position. D) High resolution volumes of the 2 ROIs registered and overlaid with low resolution overviews to show their position in context.

**Supplementary Figure 8. Resin-HiTT-targeted SBF-SEM imaging of a glomerulus ROI and a PTC-rich ROI (patient 3).** A) 3D visualization of the HiTT data acquired from the heavy metal stained, resin embedded vibratome slice. Two regions of interest (ROIs) containing a glomerulus (green) and a region rich in peritubular capillaries (yellow) were identified and segmented in the HiTT data. Two different grids were used to capture the ROIs (instead of one grid covering both glomerulus and PTC ROIs) resulting in different coloured grids. B) Low resolution overview image of the block surface acquired using SBF-SEM. The two ROIs identified in the HiTT volume are shown (green and yellow masks). C) Screenshot of the SBEMimage interface showing a grid that covers the entire imaging field. Active tiles (green) differ for each block surface exposed during cutting according to the size and position of the ROIs. The crosshair (red) shows the current stage position. D) High resolution volumes of the 2 ROIs registered and overlaid with low resolution overviews to show their position in context.

**Supplementary Figure 9. Locating and identifying immune cells via correlation of multimodal data in MoBIE (patient 1).** A) 2D snapshot of confocal fluorescence (CD16-AF647 in red and Hoechst H33342 in blue), resin HiTT and SBF-SEM data registered in MoBIE using cell nuclei as landmarks. B-D) CD16-positive immune cell in fluorescence microscopy (panel B), SBF-SEM (panel C) and overlaid fluorescence and EM data (panel D). E-G) CD16-negative immune cell in fluorescence microscopy (panel E), SBF-SEM (panel F) and overlaid fluorescence and EM data (panel G).

**Supplementary Figure 10. Locating and identifying immune cells via correlation of multimodal data in MoBIE (patient 3).** A) 2D snapshot of confocal fluorescence (CD16-AF647 in red and Hoechst H33342 in blue), resin HiTT and SBF-SEM data registered in

MoBIE using cell nuclei as landmarks. B-D) CD16-positive and CD16-negative immune cells in fluorescence microscopy (panel B), SBF-SEM (panel C) and overlaid fluorescence and EM data (panel D).

**Supplementary Figure 11. Multimodal data registration shows that immune cells identified in SBF-SEM are visible in resin HiTT and fixed hydrated HiTT.** Immune cells identified using SBF-SEM (left column) were found to be visible in resin HiTT (middle column) and fixed hydrated HiTT (right column) images.

**Supplementary Figure 12. Gallery of immune cells located and imaged using the nanopathology pipeline (patient 1).** Each intraluminal leukocyte manually segmented in 3D for downstream analysis in patient 1 is represented by a single patch. Numbering corresponds to Supplementary Table 1 rows. PLT are not shown. Confocal fluorescence images (Hoechst in blue, CD16 in red) are superimposed onto ultrastructure in greyscale Glom, glomerulus; PTC, peritubular capillary.

**Supplementary Figure 13. Gallery of immune cells located and imaged using the nanopathology pipeline (patient 2).** Each intraluminal leukocyte manually segmented in 3D for downstream analysis in patient 2 is represented by a single patch. Numbering corresponds to Supplementary Table 1 rows. PLT are not shown. Confocal fluorescence images (Hoechst in blue, CD16 in red) are superimposed onto ultrastructure in greyscale Glom, glomerulus; PTC, peritubular capillary.

**Supplementary Figure 14. Gallery of immune cells located and imaged using the nanopathology pipeline (patient 3).** Each intraluminal leukocyte manually segmented in 3D for downstream analysis in patient 3 is represented by a single patch. Numbering corresponds to Supplementary Table 1 rows. PLT are not shown. Confocal fluorescence images (Hoechst in blue, CD16 in red) are superimposed onto ultrastructure in greyscale Glom, glomerulus; PTC, peritubular capillary.

### Supplementary figures

#### Supplementary Figure 1

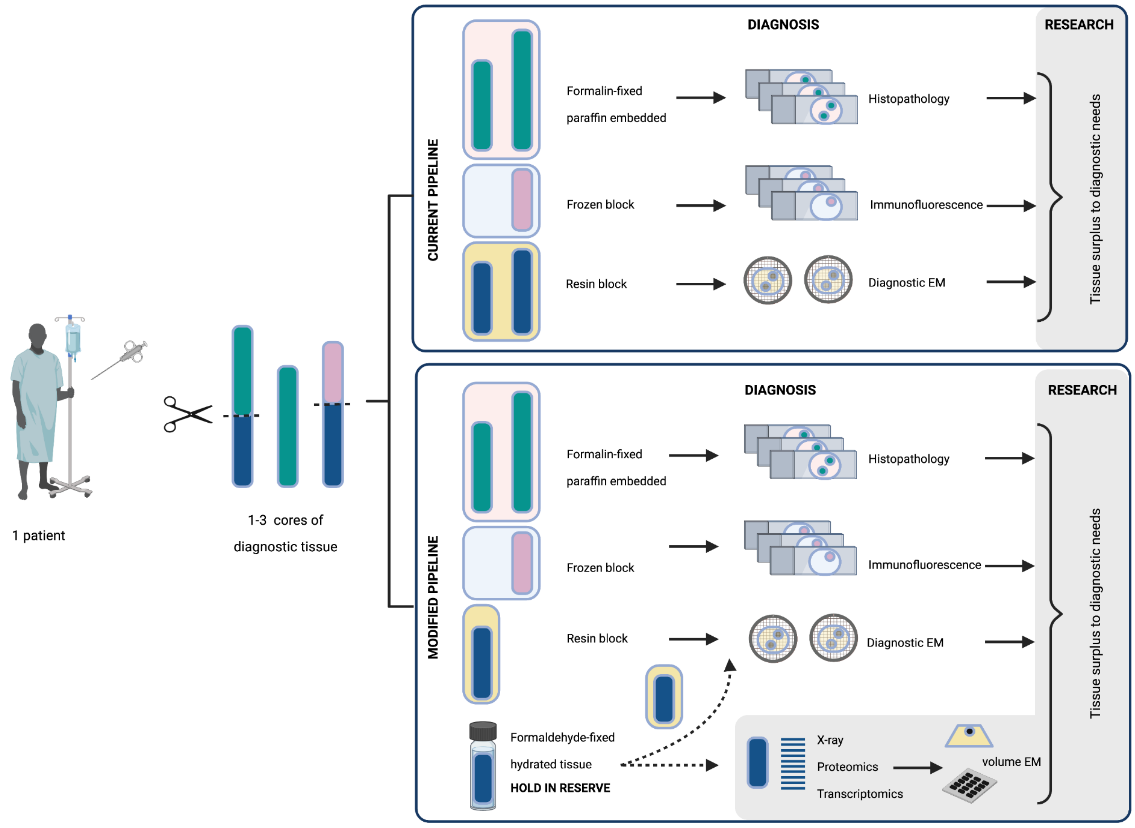

#### Supplementary Figure 2

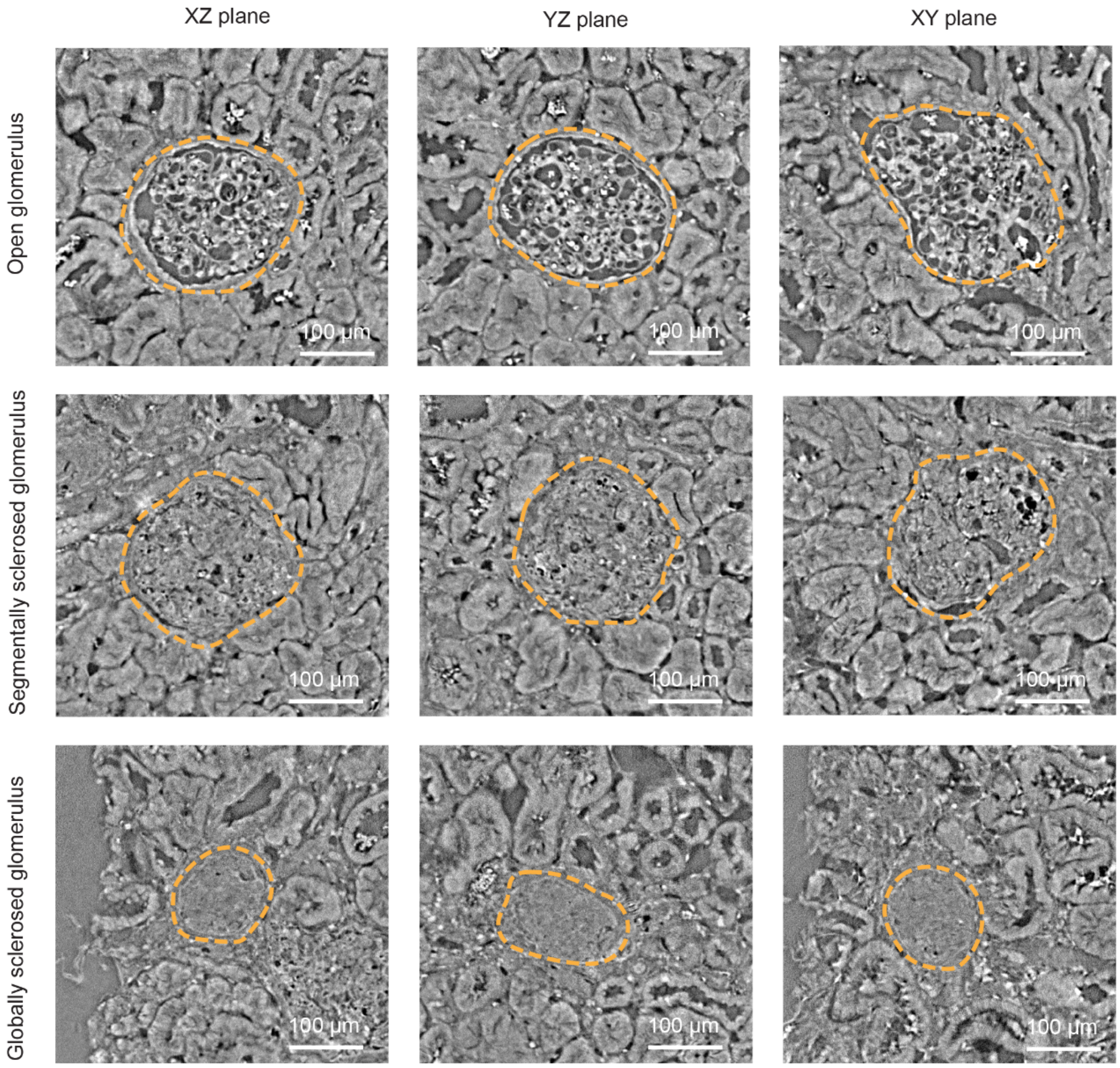

**Supplementary Figure 3**

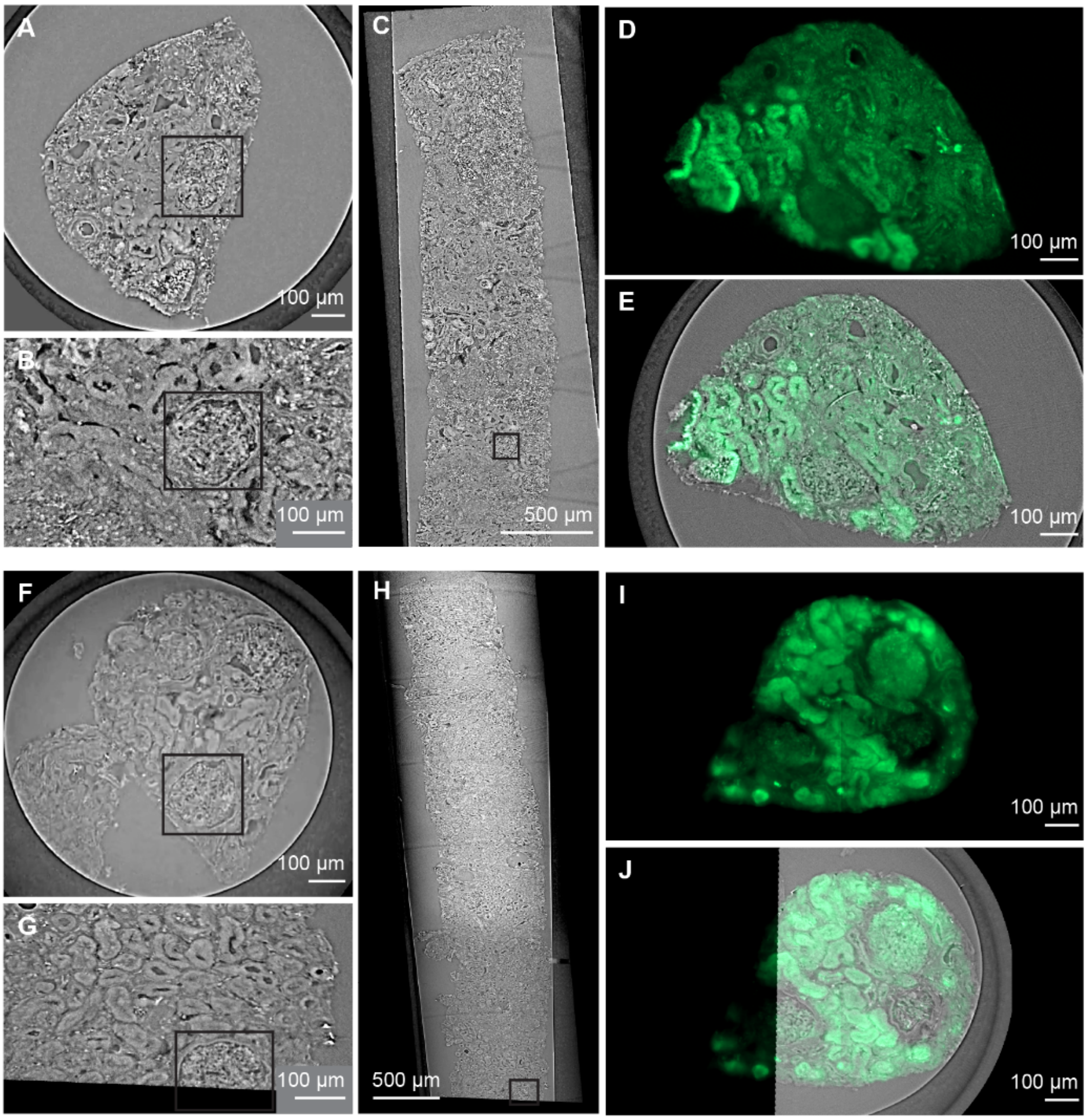

**Supplementary Figure 4**

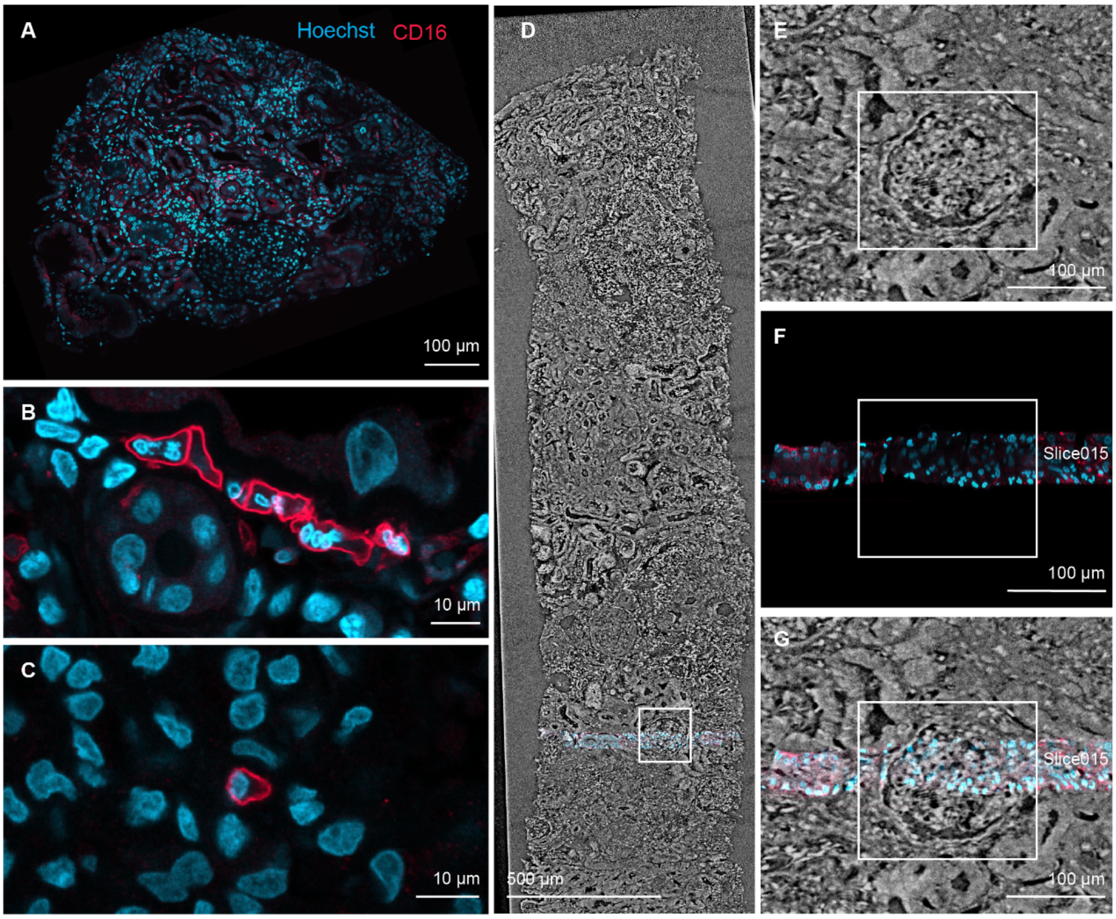

**Supplementary Figure 5**

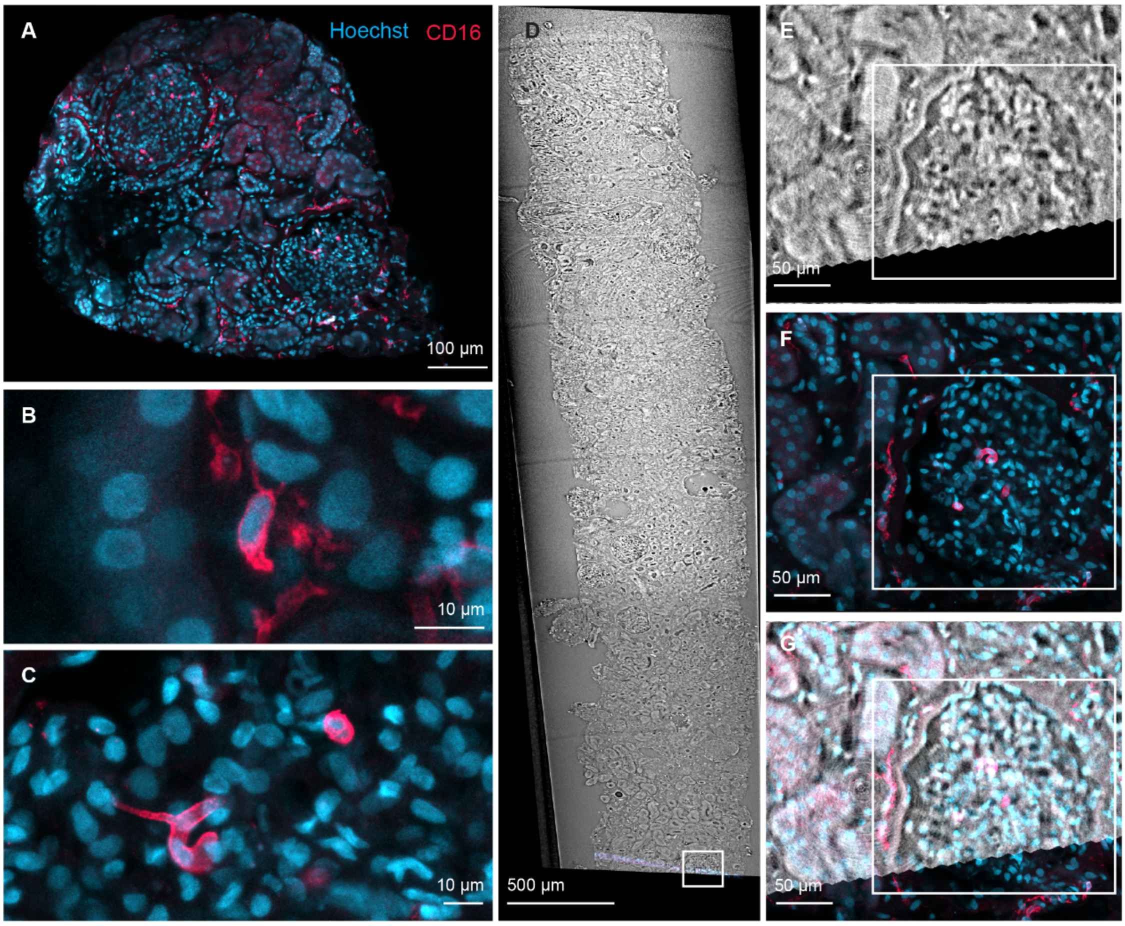

**Supplementary Figure 6**

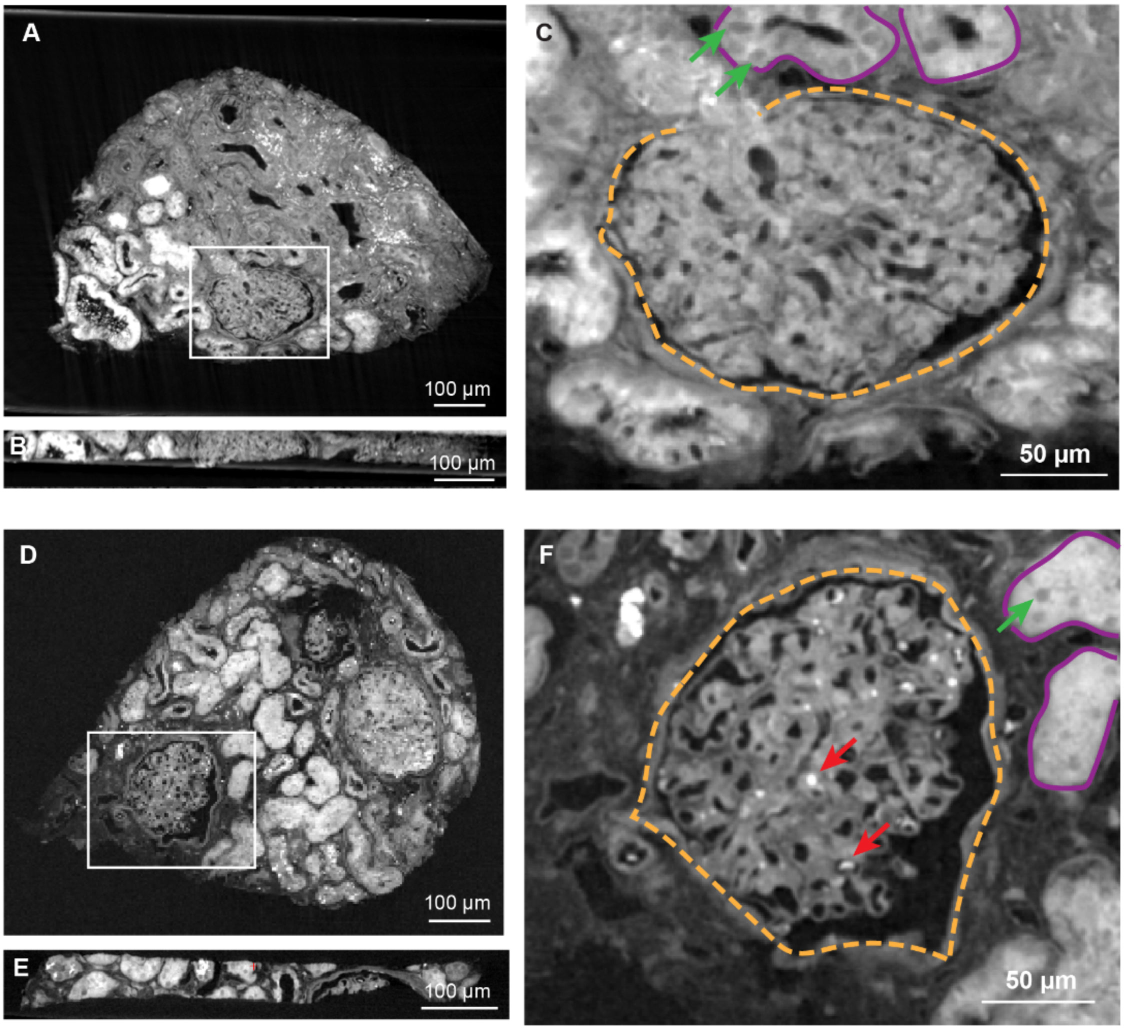

#### Supplementary Figure 7

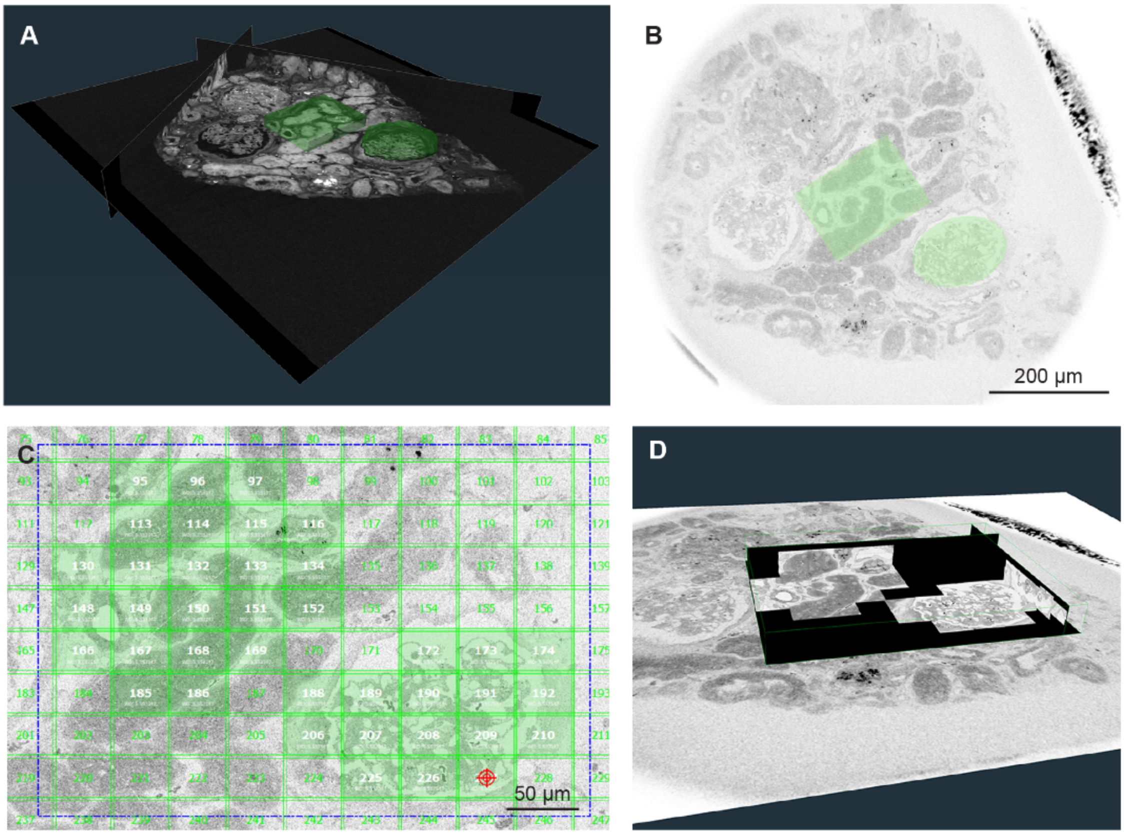

#### Supplementary Figure 8

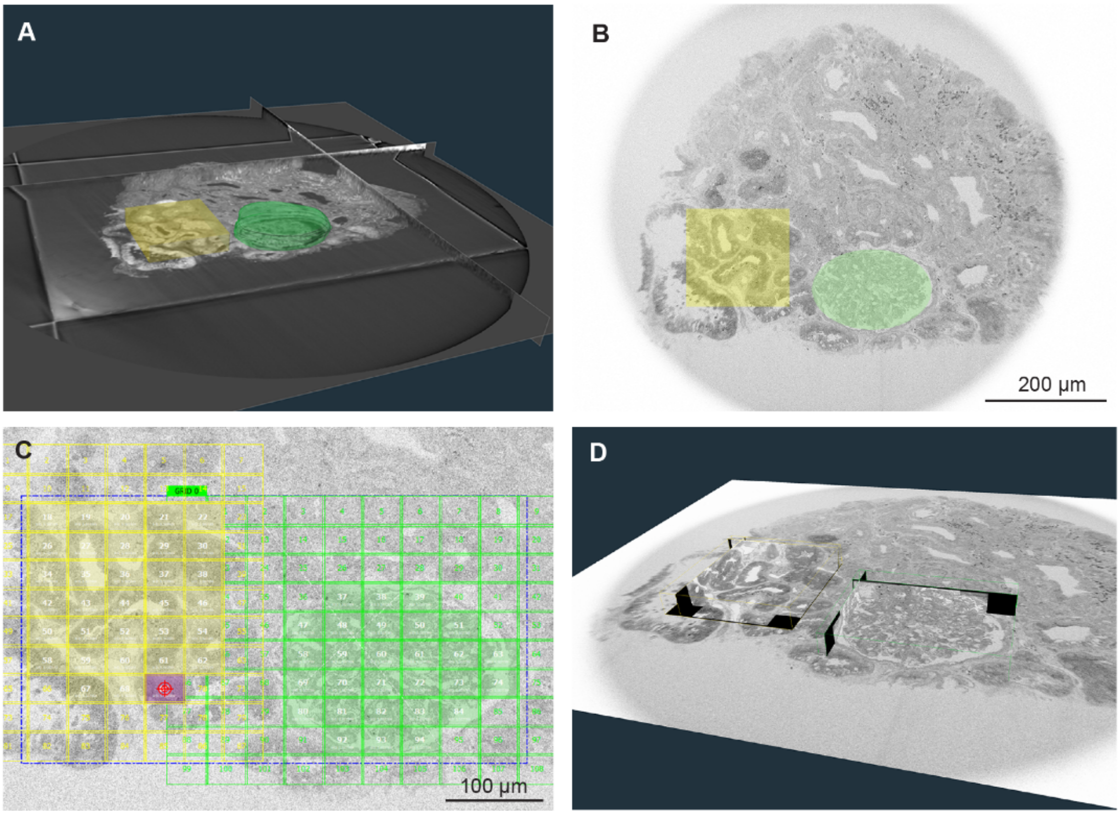

**Supplementary Figure 9**

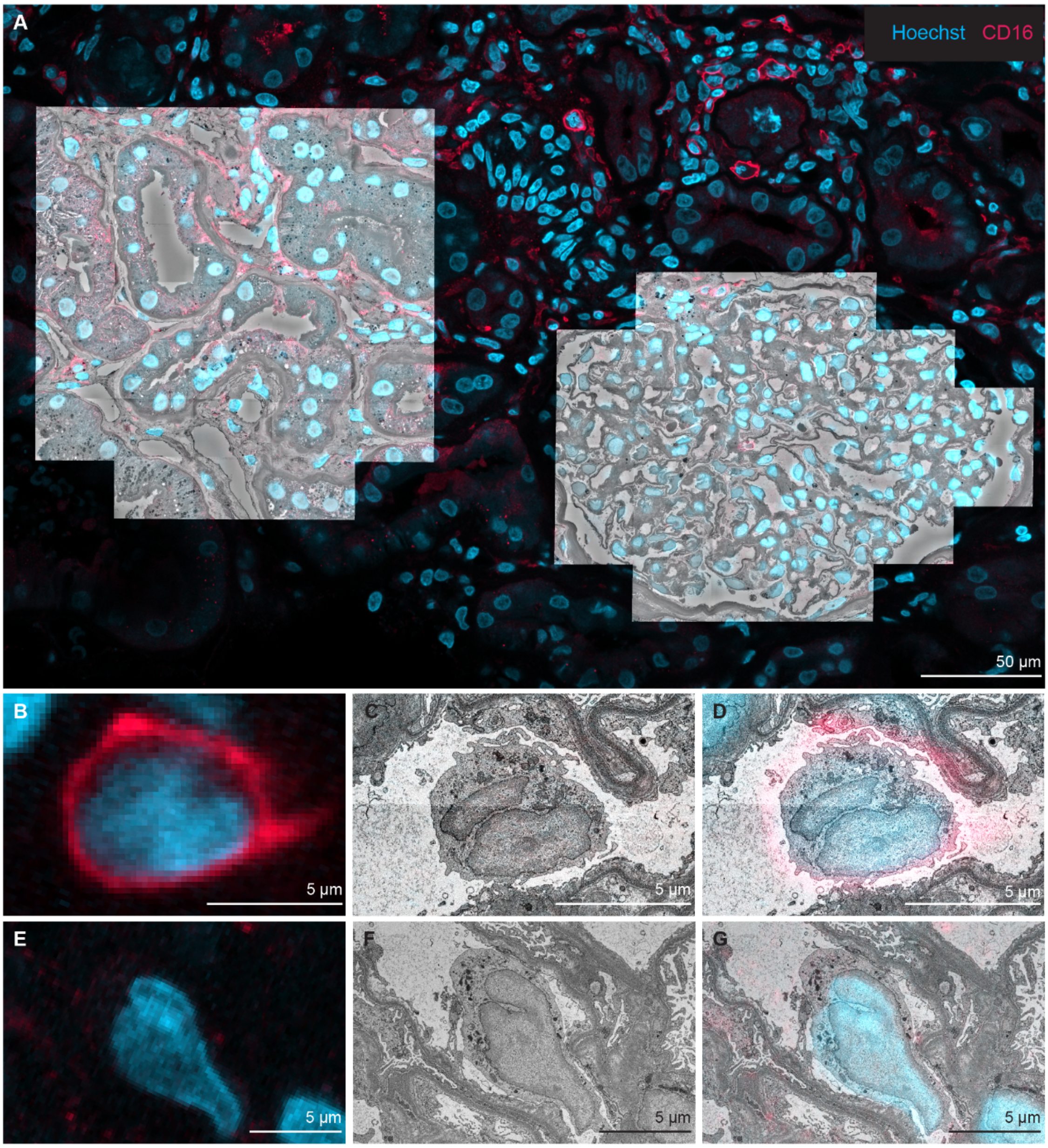

**Supplementary Figure 10**

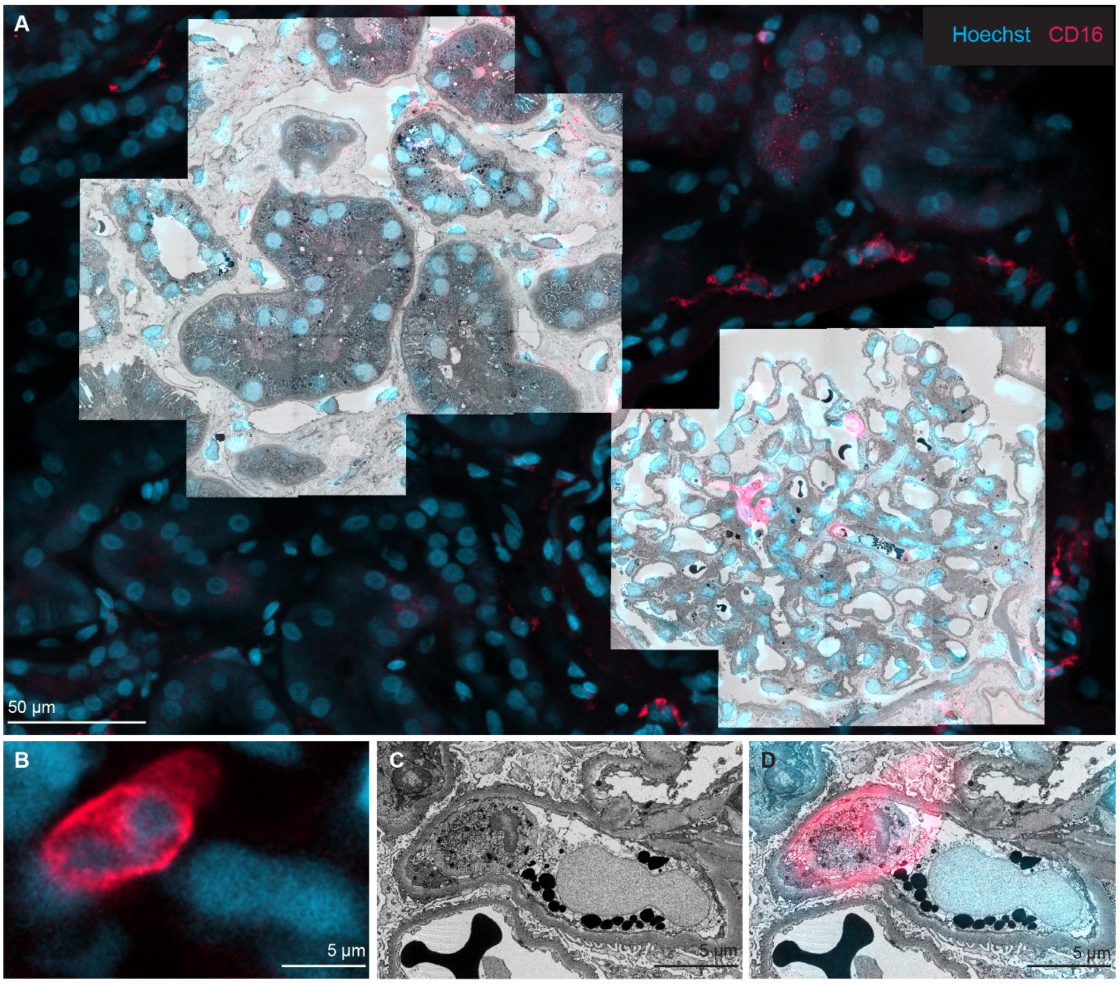

#### Supplementary Figure 11

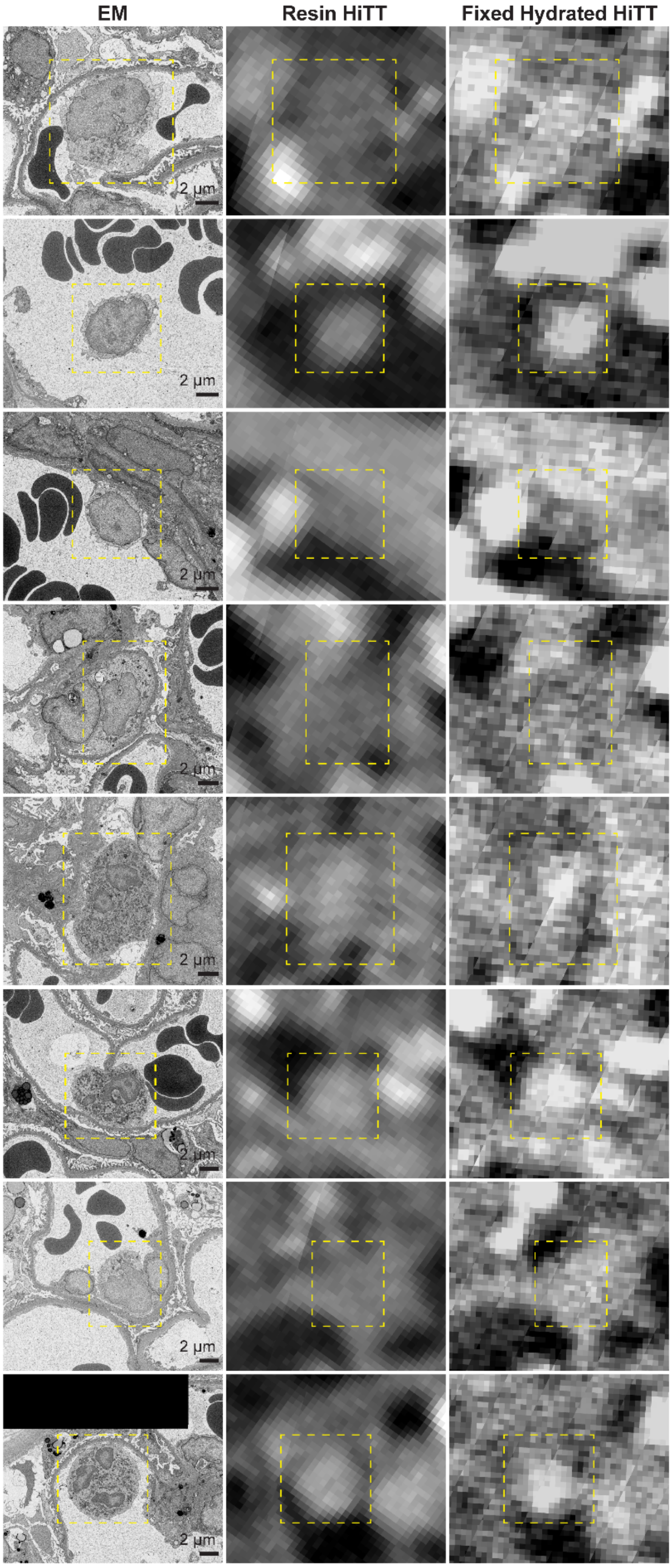

**Supplementary Figure 12**

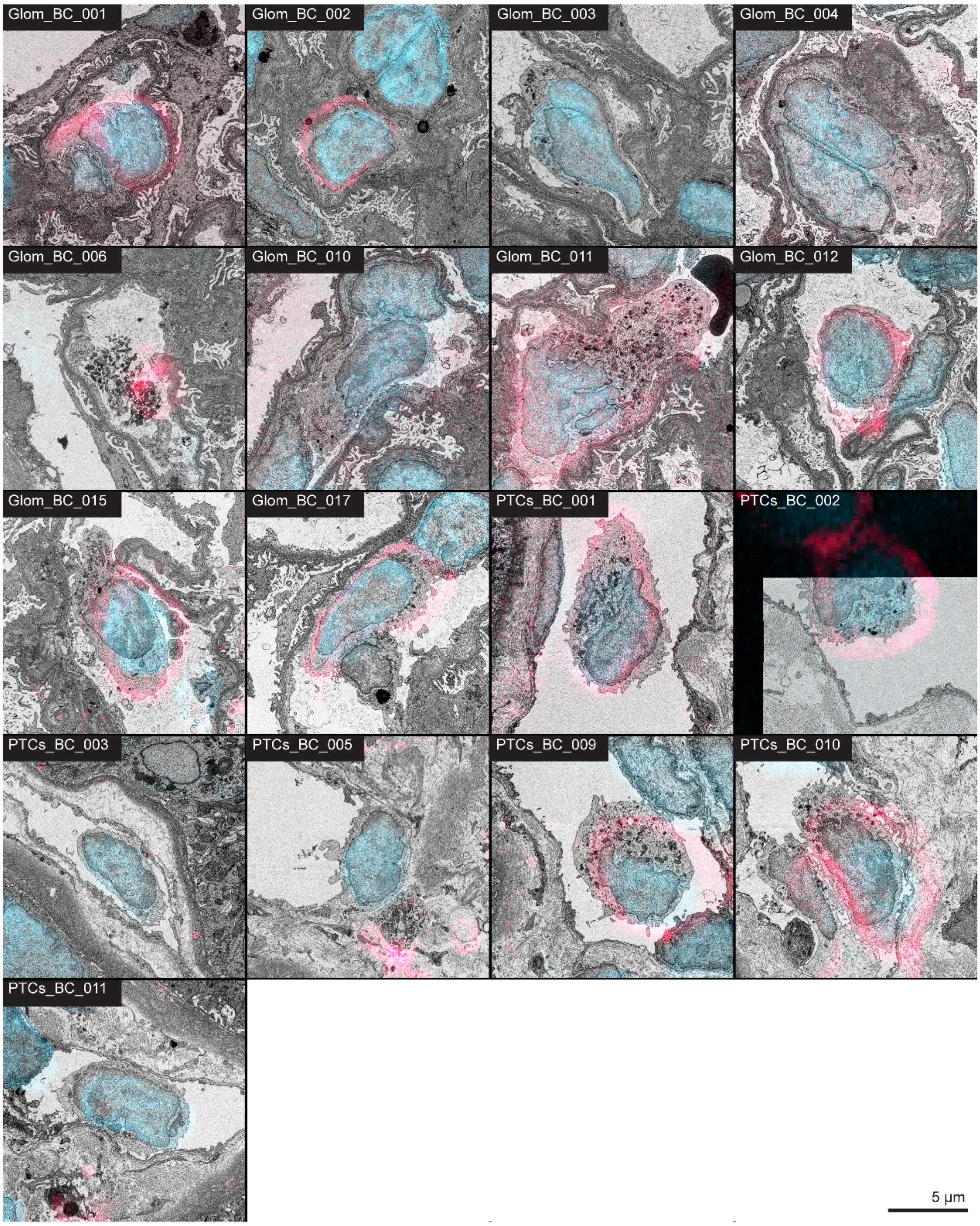

#### Supplementary Figure 13

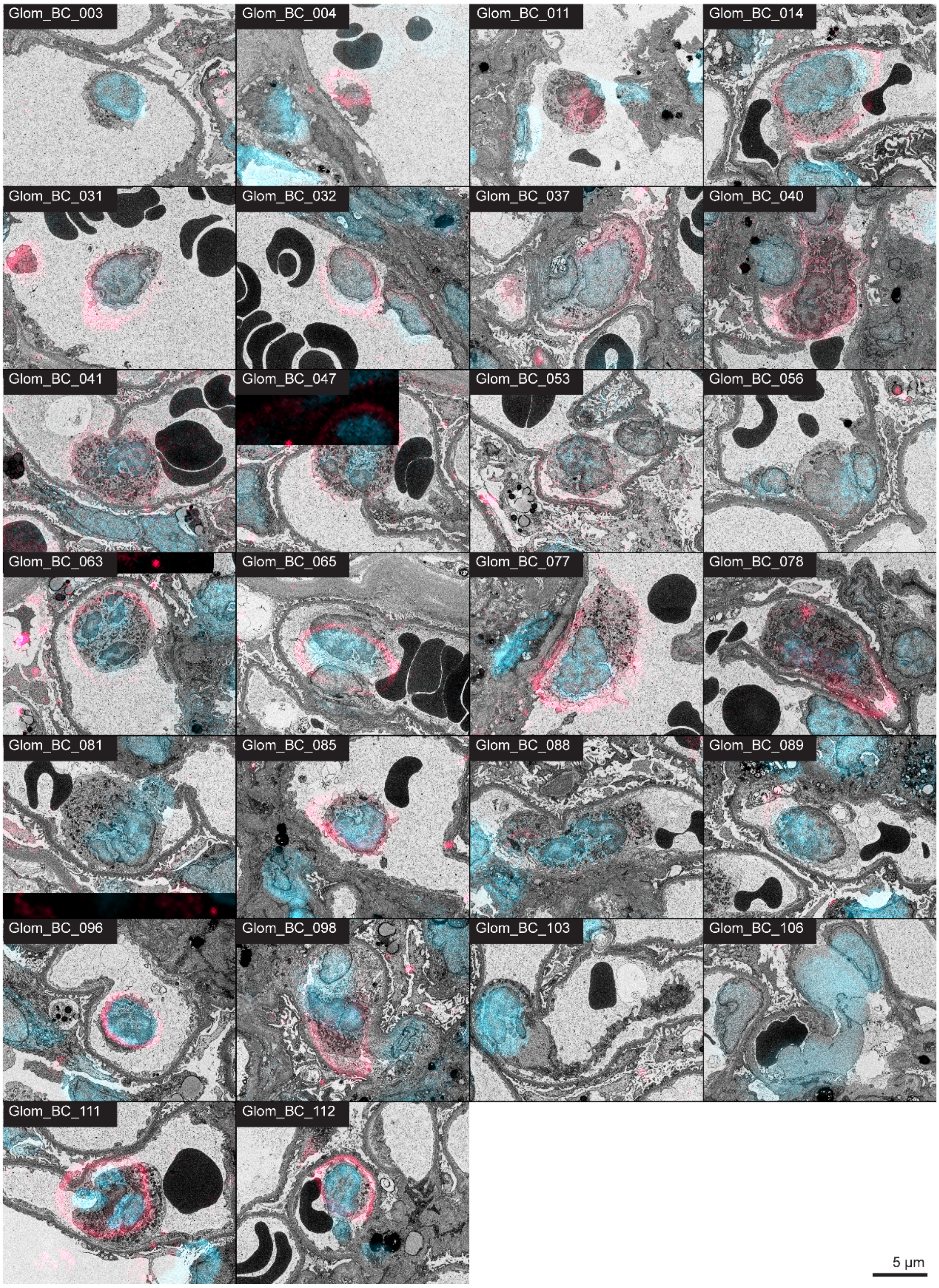

**Supplementary Figure 14**

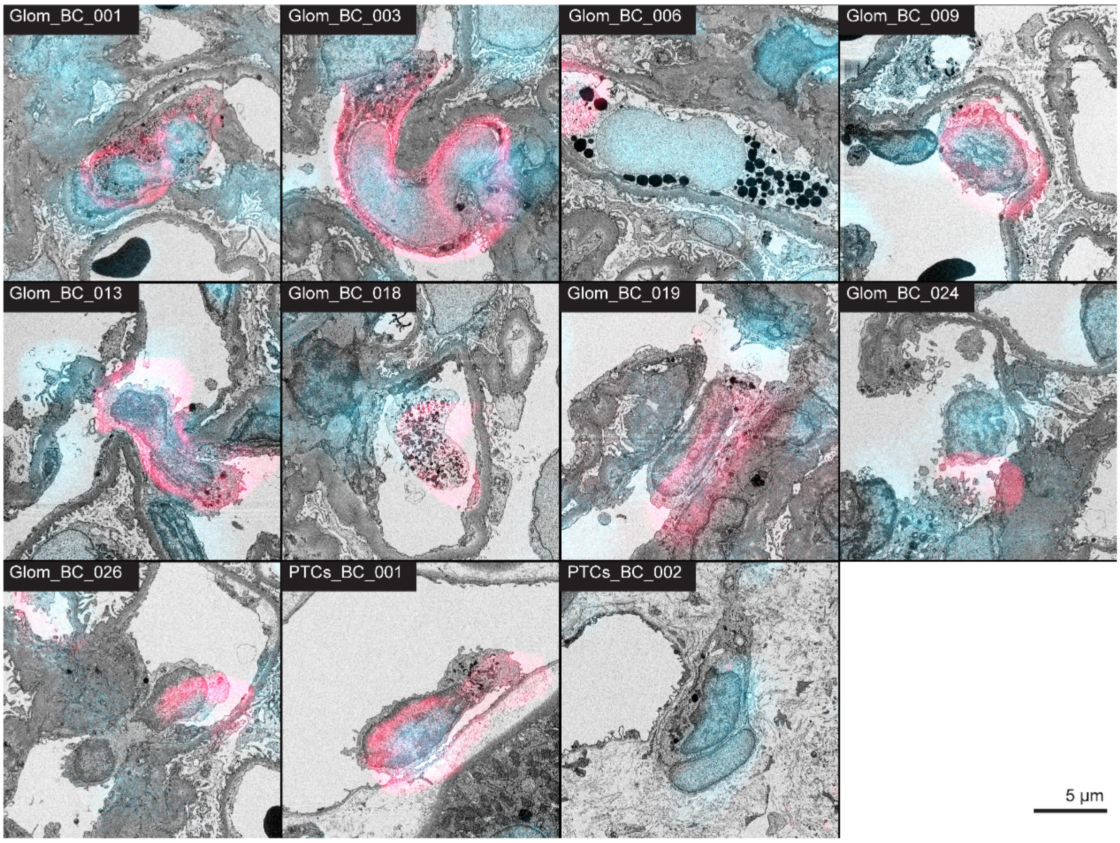

#### Supplementary Movie Legends

**Supplementary Movie 1.** Fixed hydrated HiTT of patient 1 biopsy.

**Supplementary Movie 2.** Fixed hydrated HiTT of patient 2 biopsy.

**Supplementary Movie 3.** Fixed hydrated HiTT of patient 3 biopsy.

**Supplementary Movie 4.** Movie of correlative light and electron microscopy data from patient 1 cell Glom_BC_001 in Fig.9.

**Supplementary Movie 5.** Movie of correlative light and electron microscopy data from patient 1 cell Glom_BC_012 in Fig.9.

**Supplementary Movie 6.** Movie of correlative light and electron microscopy data from patient 2 cell Glom_BC_053 in Fig.9.

**Supplementary Movie 7.** Movie of correlative light and electron microscopy data from patient 2 cell Glom_BC_103 in Fig.9.

**Supplementary Movie 8.** Movie of correlative light and electron microscopy data from patient 3 cell PTCs_BC_001 in Fig.9.

**Supplementary Movie 9.** Movie of correlative light and electron microscopy data from patient 3 cell Glom_BC_002 in Fig.9.
